# Modelling and simulation of COVID-19 propagation in a large population with specific reference to India

**DOI:** 10.1101/2020.04.30.20086306

**Authors:** Ashish Menon, Nithin K Rajendran, Anish Chandrachud, Girish Setlur

## Abstract

Deterministic mathematical models (called Compartmental models) of disease propagation such as the SIR model and its variants (MSIR, Carrier state, SEIR, SEIS, MSEIR, MSEIRS models) are used to study the propagation of COVID19 in a large population with specific reference to India.

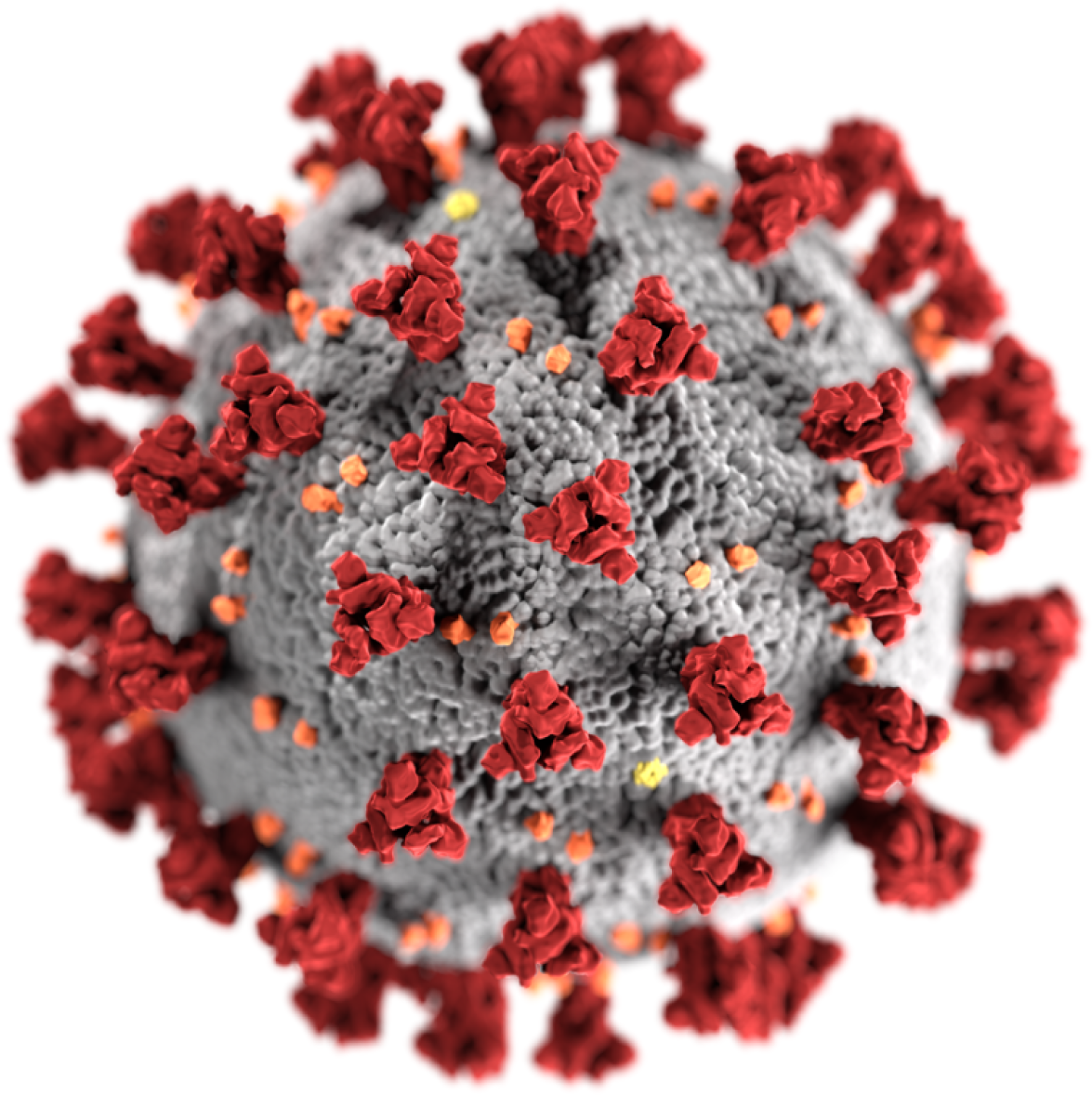

## 1 Introduction

The SIR model is one of the simplest compartmental models employed to mathematically model infectious diseases. This model comprises of three compartments: susceptible (*S*), infectious (*I*) and recovered (*R*).

The first two compartments - Susceptible and Infectious - have self-explanatory names. “Recovered” can, however, be looked at from a broader perspective. More generally, we can say that *R* consist of all the immune individuals. An immune individual can either be recovered or deceased, because neither of them can catch nor transmit the concerned disease. Each of these quantities is time-dependent, owing to the progressive nature of the disease.

Assuming a deterministic system, we model the epidemic using ordinary differential equations. By deterministic, we mean the state of each compartment at any instant in time is completely determined by the initial conditions (of the system) along with the differential equations. In simple models vital dynamics like the birth rate and death rate are omitted, while in more general models, they too are taken into account. We will look into both types of models in this paper.

## 2 SIR Model (omitting vital dynamics)

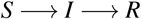

The ODEs used to model in a simple SIR model without using vital dynamics are as follows:

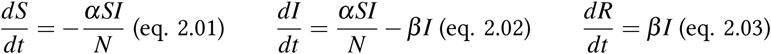

The growth rate of the disease is defined as the product of number of susceptible and infectious individuals. Here, *α* is the disease transmission rate and *β* is the recovery rate.

The nonlinear term in the above equations may be understood as follows. The rate at which the number of infected people increases with time is related to the size of the event which corresponds to infected individuals and susceptible individuals coming into close proximity with one another. The size of this event is clearly proportional to the product of the number of susceptible individuals (S) and the number of infected individuals (I). Conversely, the number of susceptible individuals falls at the same rate as they cease to be susceptible and become infected.

Equation 2.01 suggests that the number of susceptible individuals decreases with the growth rate of the disease. Since growth rate is never negative, it implies that the function *S*(*t*) is a decreasing function. Equation 2.02 suggests that the rate of increase in infectious individuals increases with the growth rate (obviously) and decreases with more individuals getting infectious. Finally, Equation 2.03 suggests the recovery (immunity) rate is directly proportional to the number of infectious individuals. This implies that *R*(*t*) is an increasing function.

Since the total population must remain constant, we have an additional equation *S*(*t*) *+ I*(*t*) *+ R*(*t*) *= N* (constant). Our objective is to develop a model for any country irrespective of their total population count. So, it makes sense to divide the above equations by their total population. This way *S*, *I* and *R* are rendered fractional values. To build up more on this idea, we will introduce some terminologies:

- Basic Reproduction Ratio: 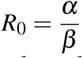. This ratio helps determine the expected number of secondary infections given a primary set (of initial conditions).
- Resolution Time: 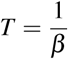. This gives a measure of the average time taken to determine the fate of an individual after contracting the disease (recovery or death).

A solution to the SIR model over the course of two weeks is shown in Figure 1.

**Figure 1:**
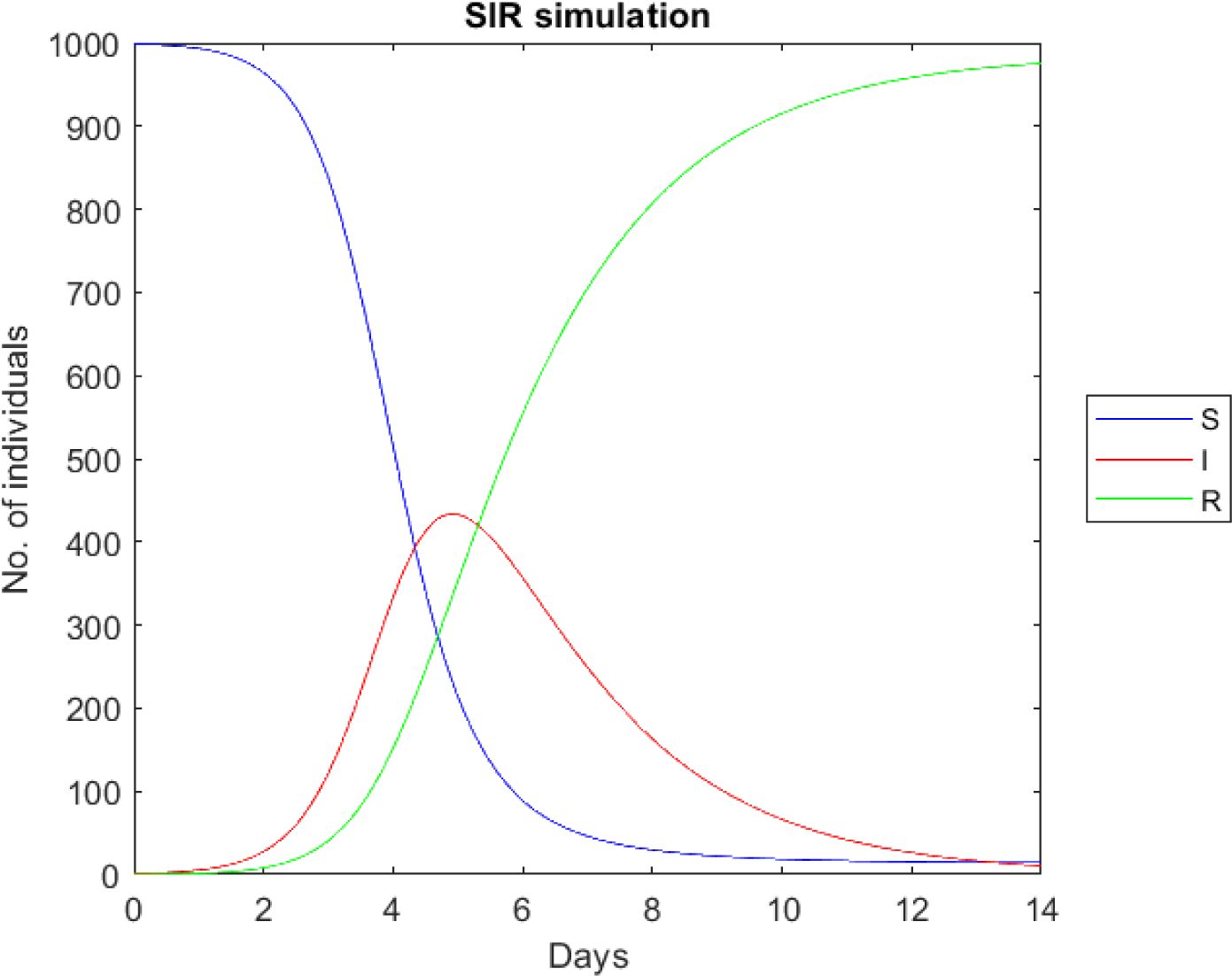
*α* = 2.18*, N* = 1000*, β* = 0.5*, S*(0) = 999*, I*(0) = 1*, R*(0) = 0

Using the parameters mentioned, it was observed that it took roughly 4.77 days for the epidemic to reach its peak value. At this time, 430 individuals were infected. It was also observed that only a meagre 14 individuals were never infected throughout the entire epidemic.

## 3 Solutions to the SIR model

### 3.1 Exact analytic solution

Until recently, only numerical methods were used to solve the SIR model. But in 2014, Harko and his coauthors derived an analytical solution.

Let us differentiate Eq. 2.01 with respect to time t. We get -

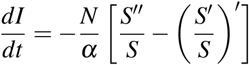

Now, eq. 2.02 transforms to,

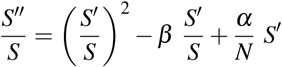

Now, eliminating I, from eq. 2.03

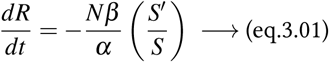

This is a simple 1st order ODE, which gives -

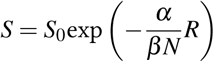

where S_0_ is a positive integration constant. Now, differentiating the above eq. we get -

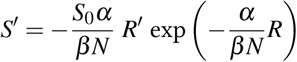

Now, if we differentiate eq 3.01 and Substitute the values of S, we get

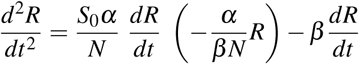

We make the following further substitutions. Set,

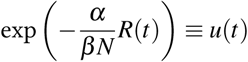

This yields,

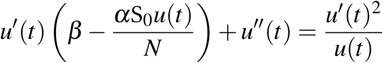

We make the following final substitution,

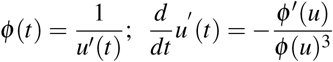

This leads to a Bernoulli type differential equation,

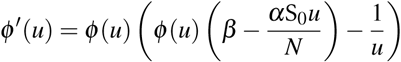

whose general solution is,

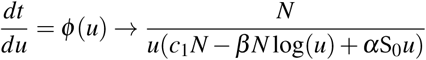

This means,

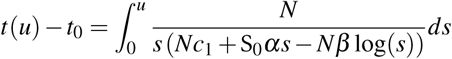

From this we may infer the time dependencies of the SIR model. However, this method is only of academic interest as modern computers obviate the need for such methods that are not easily generalizable.

### 3.2 Numerical methods

#### 3.2.1 Differential Transformation Method

Practically, it makes more sense to treat time as discrete since data is recorded not more frequently than once a day or more usually, once every few days. When this is done the following difference (as opposed to differential) equations are obtained.

A discrete version of eq. 2.01, 2.02 and 2.03 would be-

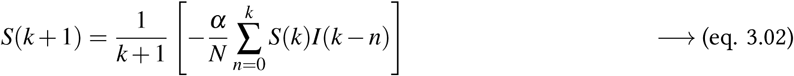

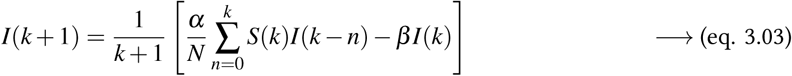

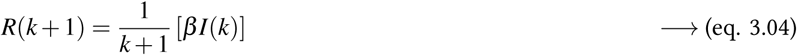

Now,

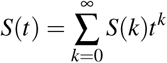

Similarly, we get analogous expressions for R and I. These difference equations may then be easily iterated on a modern computer to yield the final results.

#### 3.2.2 Variational Iteration Method

In 1999, J.H. He introduced a new method of solving coupled nonlinear ODEs called the variational iteration method (VIM). The idea behind this method is as follows.

Imagine we are called upon to solve a (likely nonlinear) ODE:

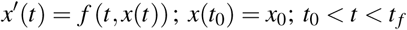

The idea is to come up with a sequence of functions *u_n_*(*t*) such that 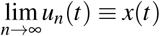. The sequence of these functions is postulated to be

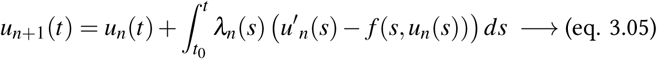

The idea is to adjust *λ*(*s*), which is called a Lagrange multiplier, in such a way that 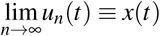 is guaranteed. Define *δu_n_*(*t*) ≡ *u_n_*(*t*) − *x*(*t*). Assuming *u_n_*(*t*) is close to *x*(*t*), we want to see how to make sure that *u_n+1_*(*t*) is even closer to *x*(*t*). Ignoring the higher powers of *δu_n_*(*t*) we get,

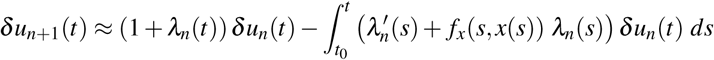

The way to make sure that *u_n+1_*(*t*) is even closer to *x*(*t*) is to ensure that *δu_n+1_*(*t*) ≈ 0. This is achieved by the following identifications.

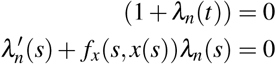

This means,

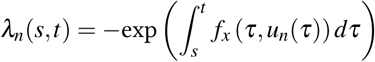

With the Lagrange multiplier fixed, we may iterate Eq. 3.04 on a computer until the desired accuracy is achieved. Before the advent of fast computers, analytical/semi-analytical methods such as these were very useful. But now we have many more options to choose from.

Applying the variational iteration method in eq. 2.01, 2.02 and 2.03; we derive the correctional functional as follows:

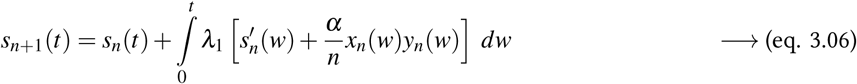

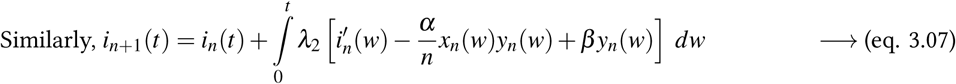

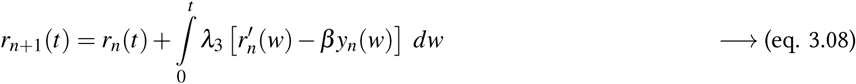

where, *λ*_1_, *λ*_2_ and *λ*_3_ are ‘Lagrange Multipliers’ and, *x_n_, y_n_* and *z_n_* are obtained as discussed above and the equations may be iterated to achieve the desired accuracy.

In this work however, we prefer to use standard numerical solvers of commercial packages such as MATLAB and Mathematica.

## 4 Adding/modifying compartments to the basic SIR model

### 4.1 SIS model

It was observed that some diseases, including common cold, did not grant any long-lasting immunity. To model such infections, the recovered (*R*) compartment had to be taken out from the SIR model.

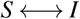

So, the only change from the ODEs describing basic SIR would be to club Equation 2.01 and 2.03.

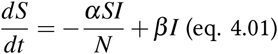

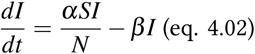

A solution to the SIS model over a course of 14 days is depicted in figure 2.

**Figure 2:**
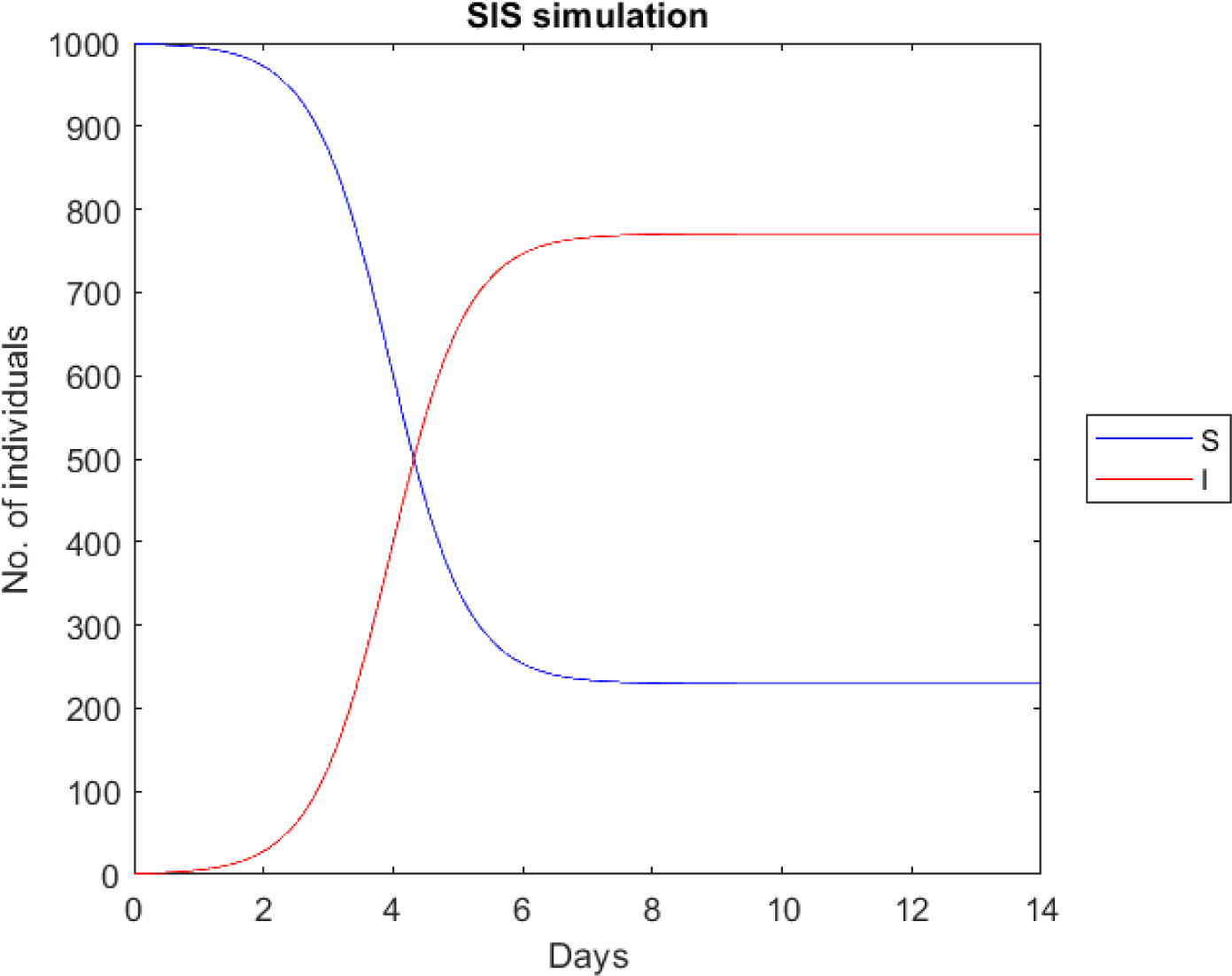
*α =* 2.18, *N =* 1000, *β =* 0.5, *s*(0) = 999, *I*(0) *=* 1

Using the parameters given above, it was observed that it took roughly 6 days for the epidemic to reach its peak value. At this time, 820 individuals were infected. It was also observed that 180 individuals were never infected throughout the entire epidemic.

### 4.2 The MSIR Model

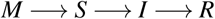

In many infections, newborns were found to be disproportionately less affected. Measles is one such infection where the babies were found to be less susceptible. This is due to a passive immunity they possess from maternal antibodies which are passed to them across the placenta.

So we require a new compartment called “Maternally-derived immunity” (*M*) to the model. Now, on loosing this passive immunity individuals in *M* compartment would transit to the *5* compartment. Gradually, a non-negative number of these individuals would further move to the *I* compartment.

However, not every new-born has this passive immunity. Only a part of them may have it (depending on whether the mothers possessed the antibodies before their birth or not). So, we assume that only a fraction *q*, of these newborns possess passive immunity and the rest do not, implies that 1 – *q* fraction of births is into the *S* compartment.

Consequently, the ODEs that are used in this model are as follows:

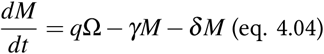

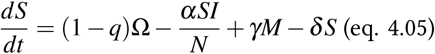

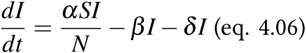

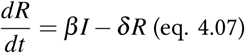

Here Ω is the birth rate and *δ* is the death rate. Since the very essence of this model is passive immunity observed in newborns, modelling without making use of vital dynamics would not have made sense. Consequently the birth rate was added to Equation 4.04 and a fraction *δ* is subtracted from all the compartments to take care of the death rate.

A simulation of a compartmental model including vital dynamics would give an idea of how extinction of the population is real possibility. An MSIR model is simulated in Fig. 3.

**Figure 3:**
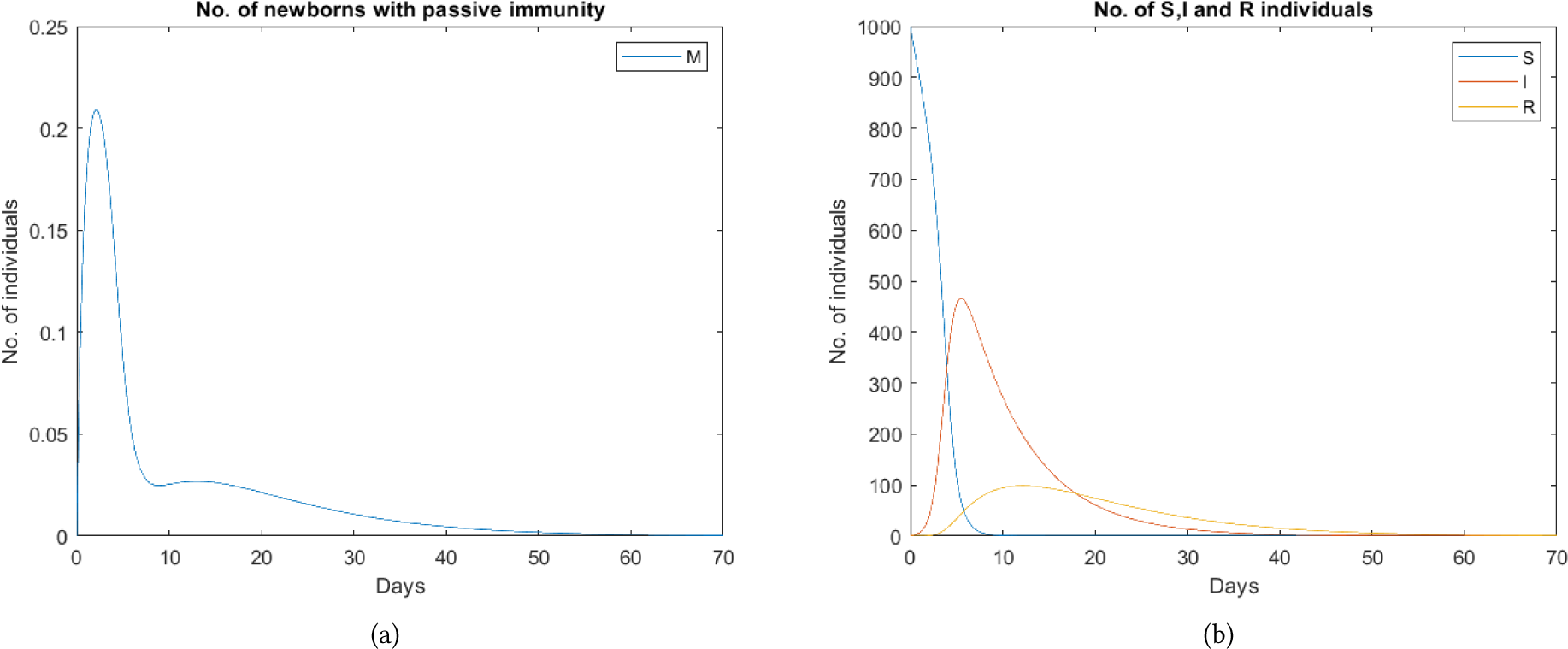
MSIR Model

Here *N =* 1000, *α =* 2.18, *β* = 0.05, Ω = 0.3, *δ =* 0.1, *q = γ =* 1. Observe that the population vanishes in roughly 70 days.

### 4.3 Carrier State

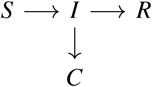

In some diseases, like tuberculosis, some individuals may never fully recover and continue to carry the infection. In due course of time, they may either fall sick again or would infect other susceptible individuals or both.

To model this infection, again a new compartment of “Carriers” (*C*) that toggle with *I* is introduced. Note that, all those cases in which the carriers both fall sick themselves and also infect other individuals is taken care of in two steps - *C → I* and *S → I*.

### 4.4 The SEIR model

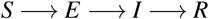

In many infectious diseases, there is an “exposed period” after the transmission of infection to susceptible individuals when they can potentially transmit the infection. This period comes before these people develop symptoms and transmit infection. So, a new compartment of “Exposed” (*E*) individuals is set up.

If the average incubation period of the infection is taken to be an exponential distribution *μ*, to incorporate this exposed period, the ODEs that have to be used are as follows:

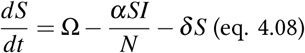

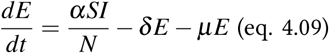

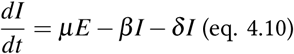

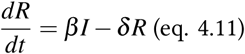

### 4.5 The SEIS model

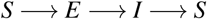

Some infections show characteristics of both the SIS and SEIR model. That is to say, they do not confer any long-lasting immunity, nor are the events of contracting the infection and becoming infectious simultaneous. Just like the SIS model, the ODEs for the SEIS model is obtained by clubbing Equation 4.08 and Equation 4.11.

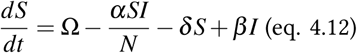

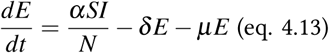

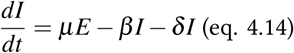

### 4.6 The MSEIR model

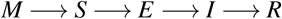

Infections showing characteristics of both the MSIR and SEIR models are modelled using an amalgamated MSEIR model. The ODEs are as follows:

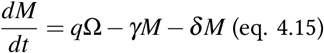

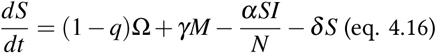

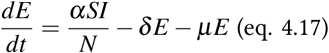

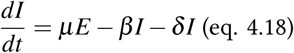

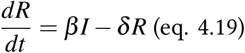

### 4.7 The MSEIRS model

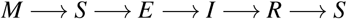

MSEIRS model is similar to the MSEIR model, except for the fact that the immunity derived by the *R* compartment is temporary and they would move back to the *S* compartment, once the immunity is lost.

## 5 Data Mining, pre-processing and modelling of COVID-19 pandemic with specific reference to India

### 5.1 Brief Overview

The crucial prerequisite for this project is the availability of data of various states in ample amount, the more the details, the better. Most often, we do not encounter clean data i.e. we usually are not able to avail data-sets in the format we desire. To be able to work with this data, it is crucial to pre-process them, which in itself is a significantly time-consuming task. The next step is to model the pandemic with simple ordinary differential equations (ODEs) using compartmental models like SIR and SIS. Various parameters will be involved in these equations. Prudent values have to be assigned to these parameters to solve the differential equations. Clues for deciding them can be obtained from the data we pre-processed. Once this is done, the evolution of all compartments with time have to be plotted to understand the nature of the pandemic and to hunt methods to ‘flatten the curve’.

The classical approach to model epidemics involves creating a set of coupled linear differential equations with multiple variables. These equations may be solved to obtain analytical solutions or can also be solved numerically using initial values and estimates(for example, using Euler’s method). Parallel computing involves solving lengthy problems by dividing them into smaller sub-problems which are solved simultaneously to obtain the complete answer to the larger problem. It is very efficient in overcoming physical constraints that prevent frequency modulation.

For simulations involving small populations of people, an efficient method is to represent each individual in the population as a string or vector of characteristic data and simulate the epidemic using computational means. This type of model is referred to as MBI (Models Based on Individuals). It can be used to include a large number of characteristics for each individual(such as age, sex, pre-existing health conditions, external factors). This method, however, is computation heavy and uses quite a lot of memory and processing time if the population being modelled is very large. For MBI, parallel processing has been used to significantly reduce computation time for models such as SIR. Simulations can be run parallely on clusters of computers making use of data from previous outbreaks. This was, in fact, used by RTI and University of Pennsylvania to model the spreading of epidemics. They used MATLAB codes and the Parallel Computing Toolbox to make models. Again, this significantly reduced processing time.

### 5.2 Data preprocessing

There are various datasets available open source for researchers to study the COVID-19 pandemic. For this project, we have chosen to work with some datasets which we feel are *authentic and exhaustive*. The [John Hopskins CSSE](https://github.com/CSSEGISandData/COVID-19) + [fixes](https://mackuba.eu/corona/overrides.json?1587060786) data set, the official figures given by the Government of India and the open source database given in www.covid19.org are some of the datasets used for collecting data on India for compiling this project. Often the datasets files are quite large and cannot be preprocessed manually. So, we have made use of technologies like MATLAB (MATrix LABoratory) to achieve this. One important dataset which we have pre-processed is the daily increase of the total confirmed cases in all indian states since the first patient was detected. The plot obtain is depicted in Figure 4.

**Figure 4.**
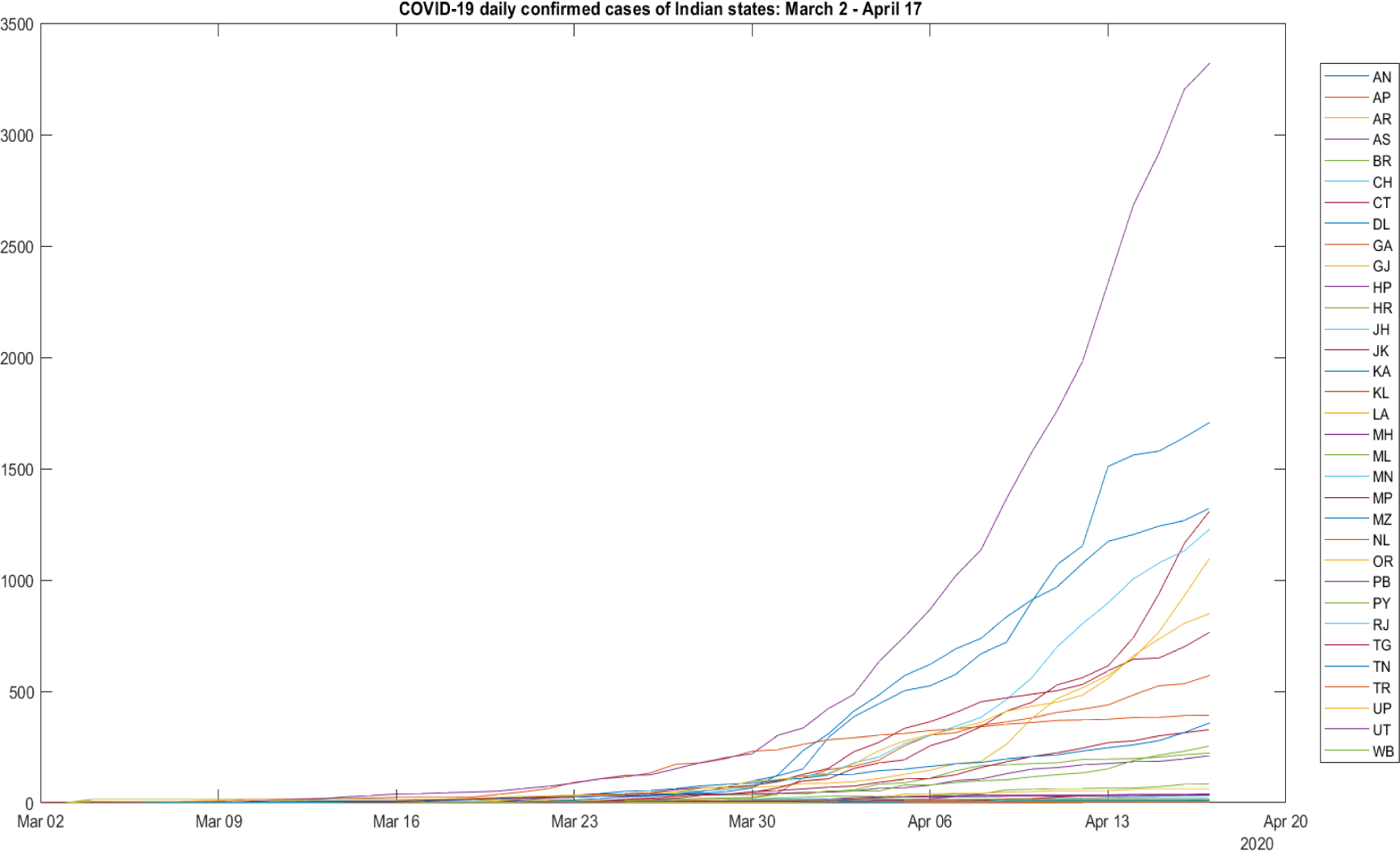

### 5.3 Modelling ODEs

#### 5.3.1 Compartments used

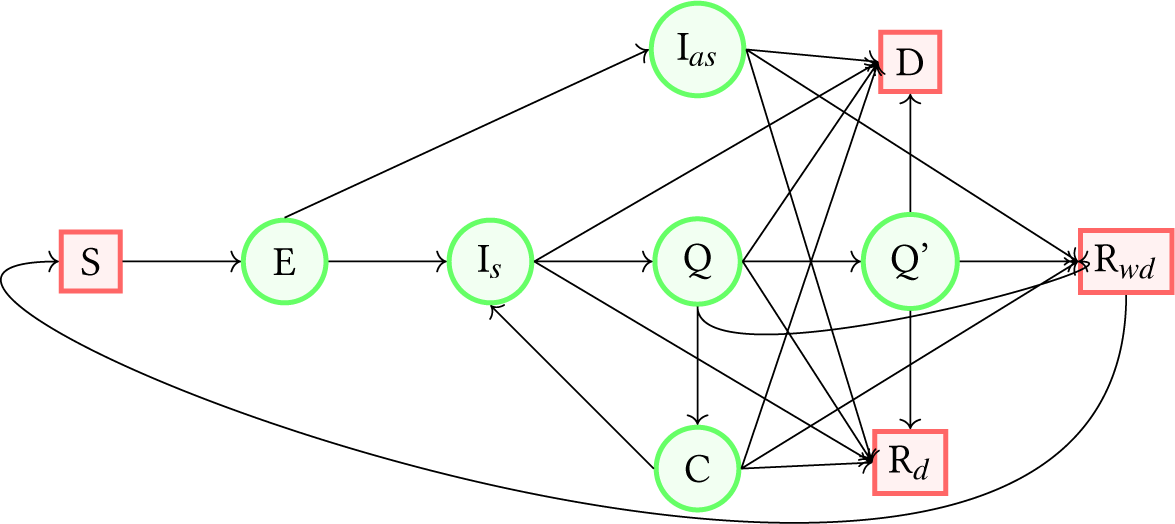

Generally speaking, we have modelled the pandemic outbreak in India using the following models:

- Susceptible (*S*): It includes all those individuals who are not infected but are susceptible to contract the disease.
- Exposed (*E*): It includes all those individuals who are exposed to infection, but are not yet infectious. With time, some of them might fall ill, some of them may not. This compartment was included keeping in mind that the incubation period of the COVID-19 pandemic [was approximated to be a significant 5-6 days by WHO](https://www.who.int/docs/default-source/coronaviruse/situation-reports/20200402-sitrep-73-covid-19.pdf?sfvrsn=5ae25bc7_2).
- Symptomatic Infected (*I*_s_): It includes all the individuals who were symptomatic and infectious. They have approached some health-care facility, but have not yet been quarantined. This compartment was brought into the picture considering concerning news from worst-affected counties like Italy and the U.S. of the hospitals and health-care centers getting filled up very fast during this pandemic. A significant portion of the infected individuals may not be quarantined in case of the health-care system collapses.
- Asymptomatic Infected (*I_as_*): It includes all those individuals who are affected but are asymptomatic before they either recover or die or get permanently disabled. This compartment was formalised keeping after reports of various [*authentic* studies](https://www.buzzfeednews.com/article/stephaniemlee/coronavirus-asymptomatic-silent-carrier-spread-contagious) claiming that around 30-40 percent of the infected individuals remain asymptomatic.
- Quarantined (*Q*): It includes all those individuals who are currently kept under quarantine in a health-care facility.
- ICU (*Q′*): It includes the quarantined patients who had to be moved to Intensive Care Unit after their condition worsened.
- Carrier (*C*): This compartment includes those individuals who have left quarantine after being tested negative, but actually have not fully healed. So, they possess the ability to infect other susceptible individuals. They eventually either fall sick again or recover from the disease. This compartment was introduced after multiple cases of re-infection being reported from countries like South Korea, China and Japan. While we fully do not understand the reason for these reports yet, it is safe to assume that one of the following happened:

1. Due to medical inefficiency (inaccurate result)
2. Due to loss of immunity after recovery and subsequent re-infection.
- To account for the first possibility, we have introduced the carrier compartment. To account for the second possibility, we have kept the transition from the recovered (*R*) compartment to susceptible (*S*) compartment a possibility, while writing the ODEs.
- Recovered without disability (*R_wd_*): In this compartment, we have kept all those individuals who have recovered from the infection without any disability and can no longer infect any other individual.
- Deceased (*D*): This compartment includes all the individuals who have fell victims to the pandemic.
- Recovered with disability (*R_d_*): In this compartment, we have kept all those individuals who have recovered from the infection, can no longer infect anyone else, but have been permanently disabled post recovery.

#### 5.3.2 Differential equations formulated

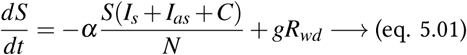

where *α* is the disease transmission rate and *g* is the rate at which a fraction of recovered individuals lose their immunity

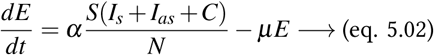

if the average incubation period is taken to be an exponential distribution *μ*.

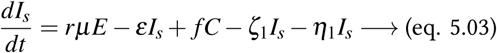

where *f* is rate at which a fraction of carriers gets re-infected. Here 0 ≤ *r* ≤ 1 is a number which gives a measure of how many individuals in the exposed compartment moves of *I_s_* compartment, rather than the *I_as_* one. Here *ε*, *ζ*_1_ and *η*_1_ are the rates at which infected individuals get quarantined, deceased and disabled respectively.

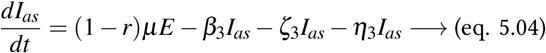

where *β*_3_*, ζ*_3_ and *η*_3_ are the recovery rate, death rate and disability rate of asymptomatic individuals, respectively.

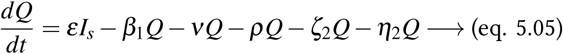

where *ν*, *ρ*, *ζ*_2_ and *η*_2_ are the rates at which the quarantined individuals go to carrier state, ICU, deceased and disabled compartments respectively. *β*_1_ is the recovery rate for quarantined individuals.

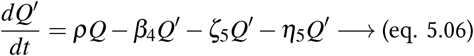

where *β*_4_, *ζ*_5_ and *η*_5_ are the rate with which individuals in ICU recover, die and get disabled, respectively.

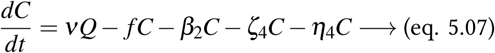

where *β*_2_, *ζ*_4_ and *η*_4_ are the rate with which carrier individuals recover, die and get disabled silently, respectively.

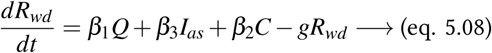

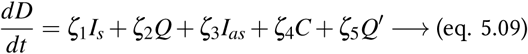

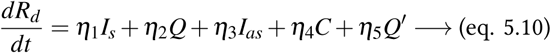

#### 5.3.3 Choice of parametric values

The parameters have been set to the following values in per unit days to simulate a typical outbreak on an epidemic using the model that has been discussed above. The choice of some of the parameters is on basis of the study [INDSCI-SIM](https://indscicov.in/for-scientists-healthcare-professionals/mathematical-modelling/indscisim/?fbclid=IwAR21o9WgQoL0yZIgI4JOWznV7f7XlPrW9GOuu9WgT823TCqoPtD1hCPlFmM) (described in a later section).

| Values of parameters |  |  |  |
| --- | --- | --- | --- |
| Parameter | Value | Parameter | Value |
| $N$ | 1 | $\alpha$ | 0.42 |
| $\beta_1$ | 0.5 | $\zeta_1$ | 0.25 |
| $\beta_2$ | 0.1458 | $\zeta_2$ | 0.2 |
| $\beta_3$ | 0.1458 | $\zeta_3$ | 0.1 |
| $\beta_4$ | 0.05 | $\zeta_4$ | 0.05 |
| $\mu$ | 0.5 | $\zeta_5$ | 0.2 |
| $\nu$ | 0.05 | $\eta_1$ | 0.1 |
| $\varepsilon$ | 0.5 | $\eta_2$ | 0.1 |
| $r$ | 0.3 | $\eta_3$ | 0.05 |
| $g$ | 0.0001 | $\eta_4$ | 0.01 |
| $f$ | 0.2 | $\eta_5$ | 0.1 |
| $\rho$ | 0.01 | | |

#### 5.3.4 Result obtained

After solving the highly-coupled differential equations given in section 5.3.2, the plot for the various compartments that was obtained is depicted in Figure 5.

**Figure 5:**
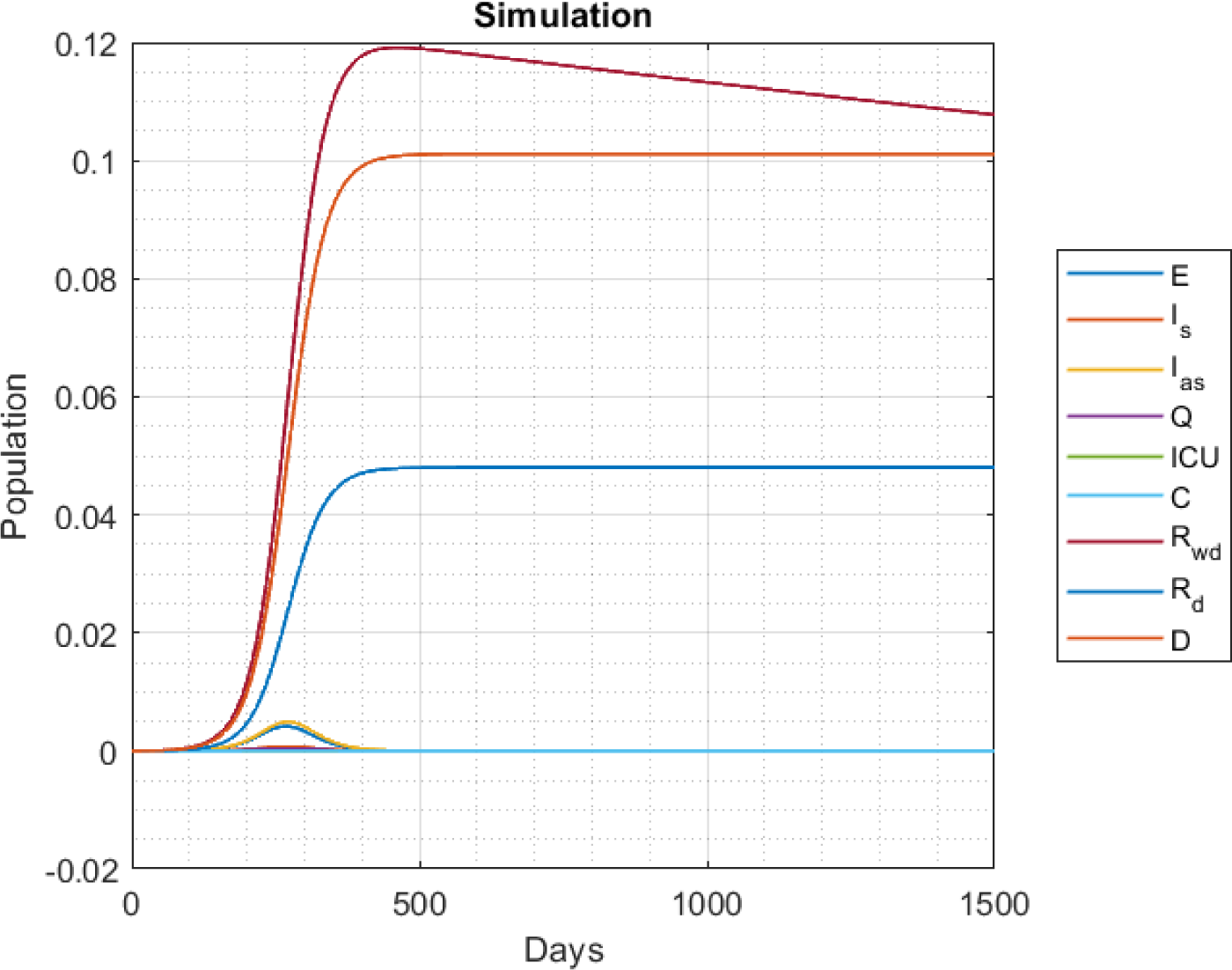
Initial conditions: S(0) = 1 − 1 × 10^−5^, E(0) = 1 × 10^−5^, others = 0

### 5.4 Effect of lock down

In this section, we study how imposing lock downs will affect the plot in figure 5. *α* denotes the number of interactions per-capita. So a decrease in alpha would mean a lower rate of contact between infected and susceptible individuals, as would be the case in a lock down. So, to simulate lock downs, we have decreased the value of the parameter *α* to 0.14, for the overall period of lock down. The simulations of no lock down, lock down for days 60-90 and lock down for days 150-200 on the total active and deceased cases are depicted in Fig. 6 and 7.

**Figure 6:**
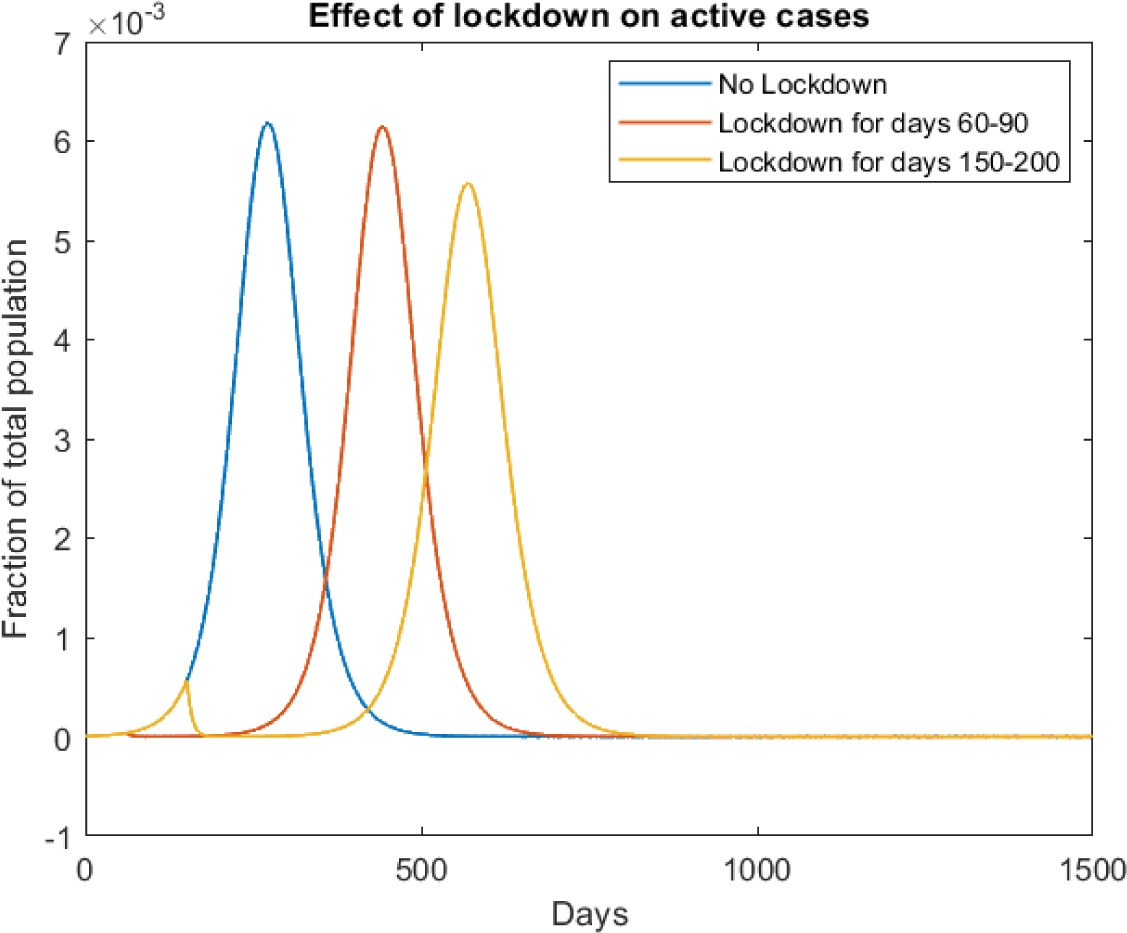
Effect on net active cases: *I_s_ + I_as_ + Q + Q′ + C*

**Figure 7:**
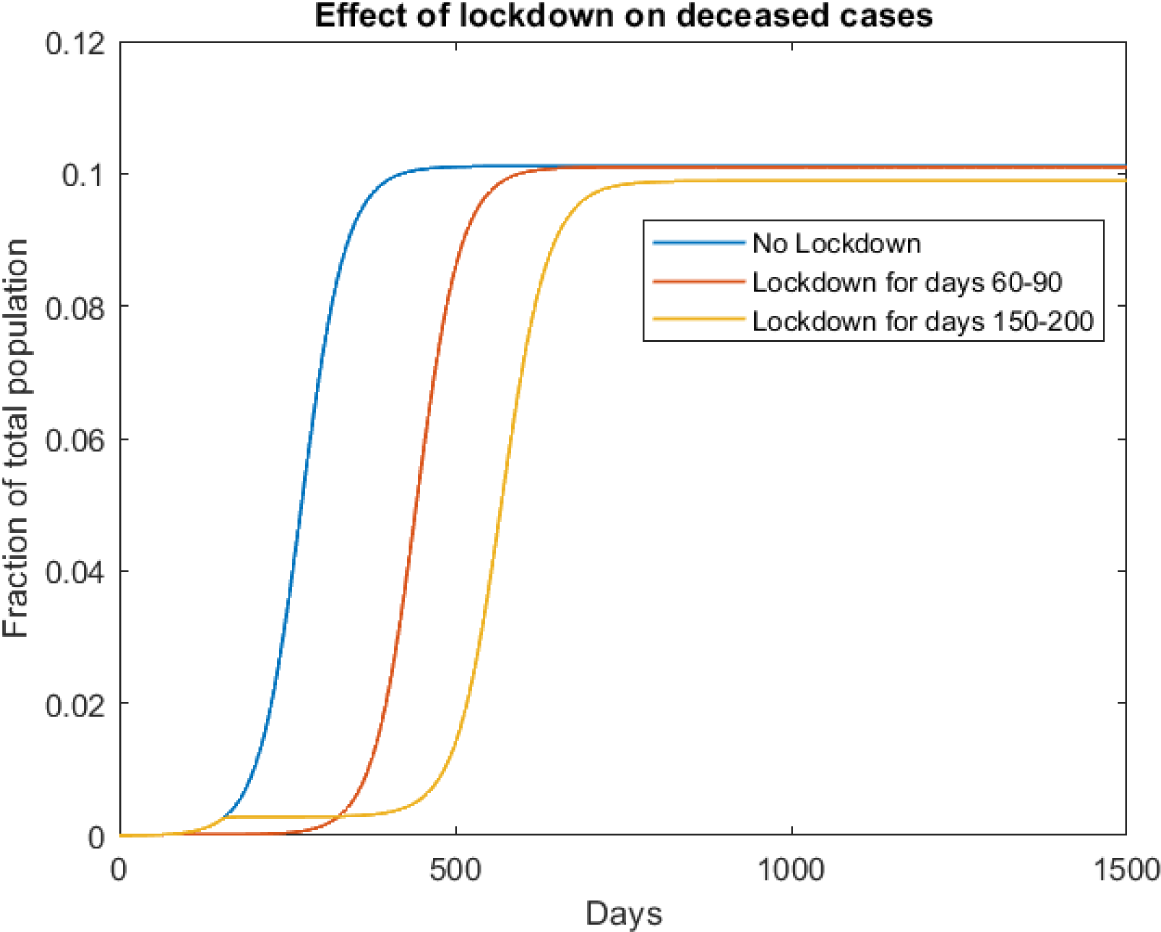
Effect on *D* compartment

It turns out that just locking down, will only delay the peak of the epidemic. It seems like a periodic lock down is actually allowing the pandemic to infect individuals in installments. Fig. 8, which is a simulation of when lock downs are imposed for both days 150-200 and days 500-550, may make this idea more clear. The total cases remain the same.

**Figure 8:**
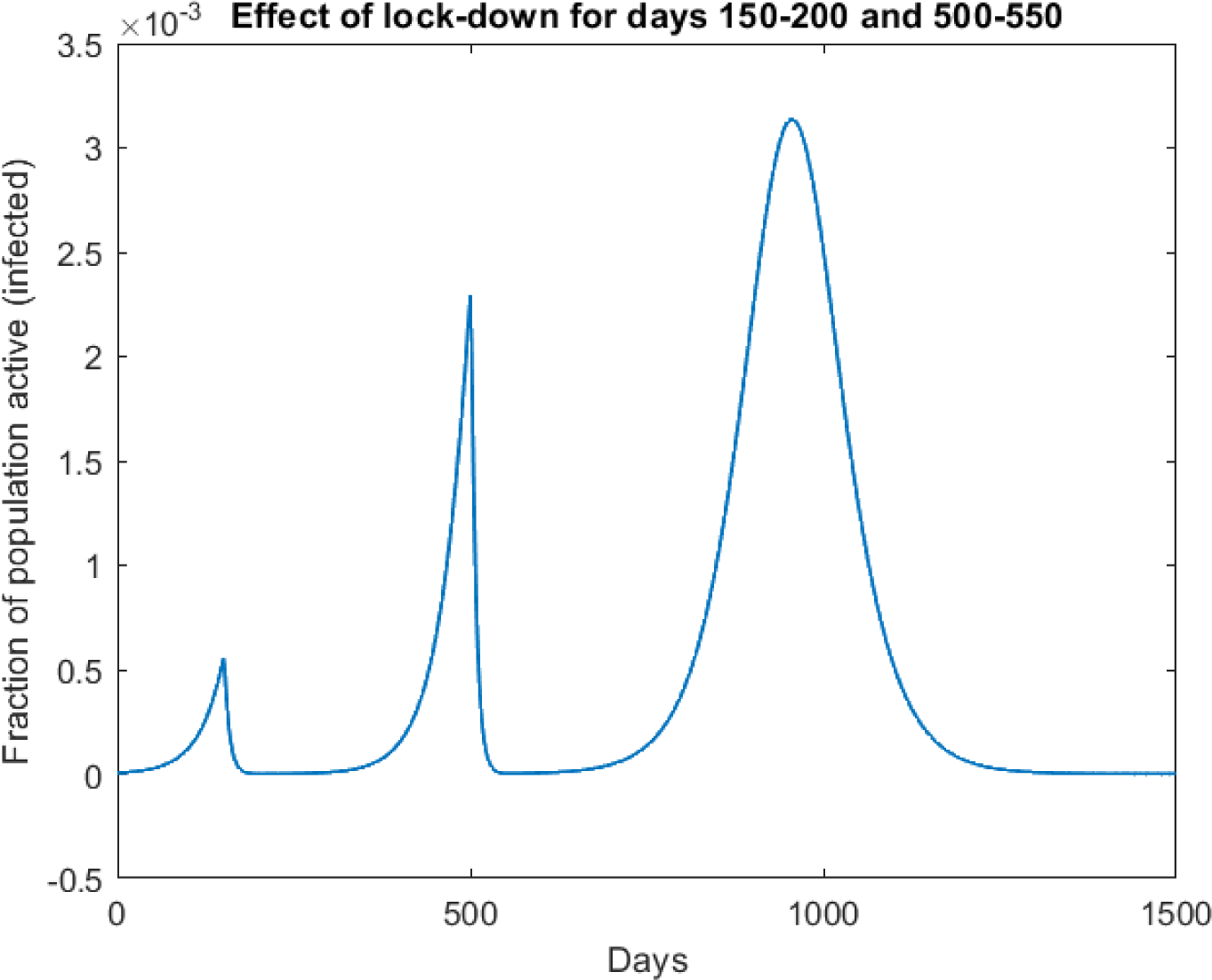
lock down for both days 150-200 and 500-550

**Assumptions made:**

- The entire system under consideration is locked down simultaneously at the point of enforcement.
- At least one individual remains infectious after the lock down is relaxed.
- No violations of lock down occur during the entire period of lock down.
- No migration from other cities/systems occurs to the system under consideration.

#### Recommendations for public policy

Understanding how lock downs affect the epidemic, the government can make use of this vulnerability in the spread of the pandemic by imposing lock downs at specific times, to slow the spread of the disease. Subsequently, effective policies should be enacted to get the situation under control and set up medical facilities, to prepare for the surge in infections that is to follow the lifting of lock down. The effect of setting up medical facilities is studied in a later subsection.

Now, these lock downs can be executed in various fashions. One model is that of continuous lock down, like the ones depicted above. But as proved above, they just delay the onset of the disease. To completely eradicate the disease, the economy (nation) would have to lock down theoretically for an impractically long time (see later). This is not possible due to the fragile nature of the Indian economy.

#### 5.4.1 Periodic lock down

In this strategy, the economy is shut down and re-opened at regular intervals to allow the economy to recover, though minimally, in the time window allotted. One strategy is the 7+5 strategy, wherein lock down is imposed (after a threshold time) for 7 days at length. Once this duration is over, it is reopened for 5 days, before going to the next lock down for 7 days again. And this process is repeated. The simulation for the 7+5 strategy on our compartmental model after the first 150 days is depicted in Fig.9.

**Figure 9:**
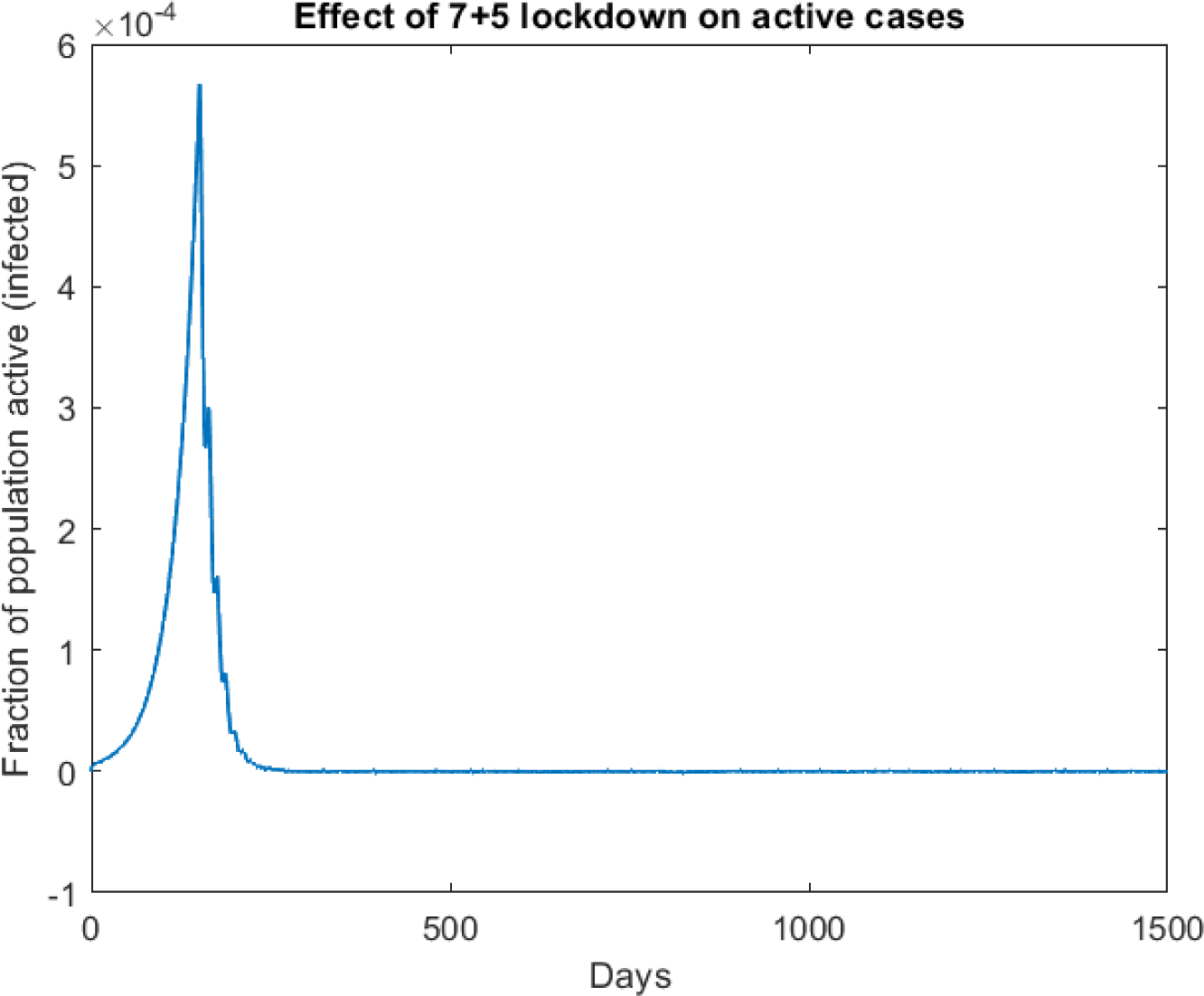
7+5 lock down from 150 days to eternity

From the Fig. 9, it seems that periodic lock down will in the long run, eradicate the disease. This is true, only if the lock down is continued to eternity, which again is impractical. In real world scenarios, the economy would have to be re-opened after some time. Fig. 10 shows what happens when you relax the periodic lock down measures after 250 days.

As evident from the Fig.10, once the periodic lock down is relaxed, the second wave of the epidemic returns, thereby making this model too unfit for use alone.

**Figure 10:**
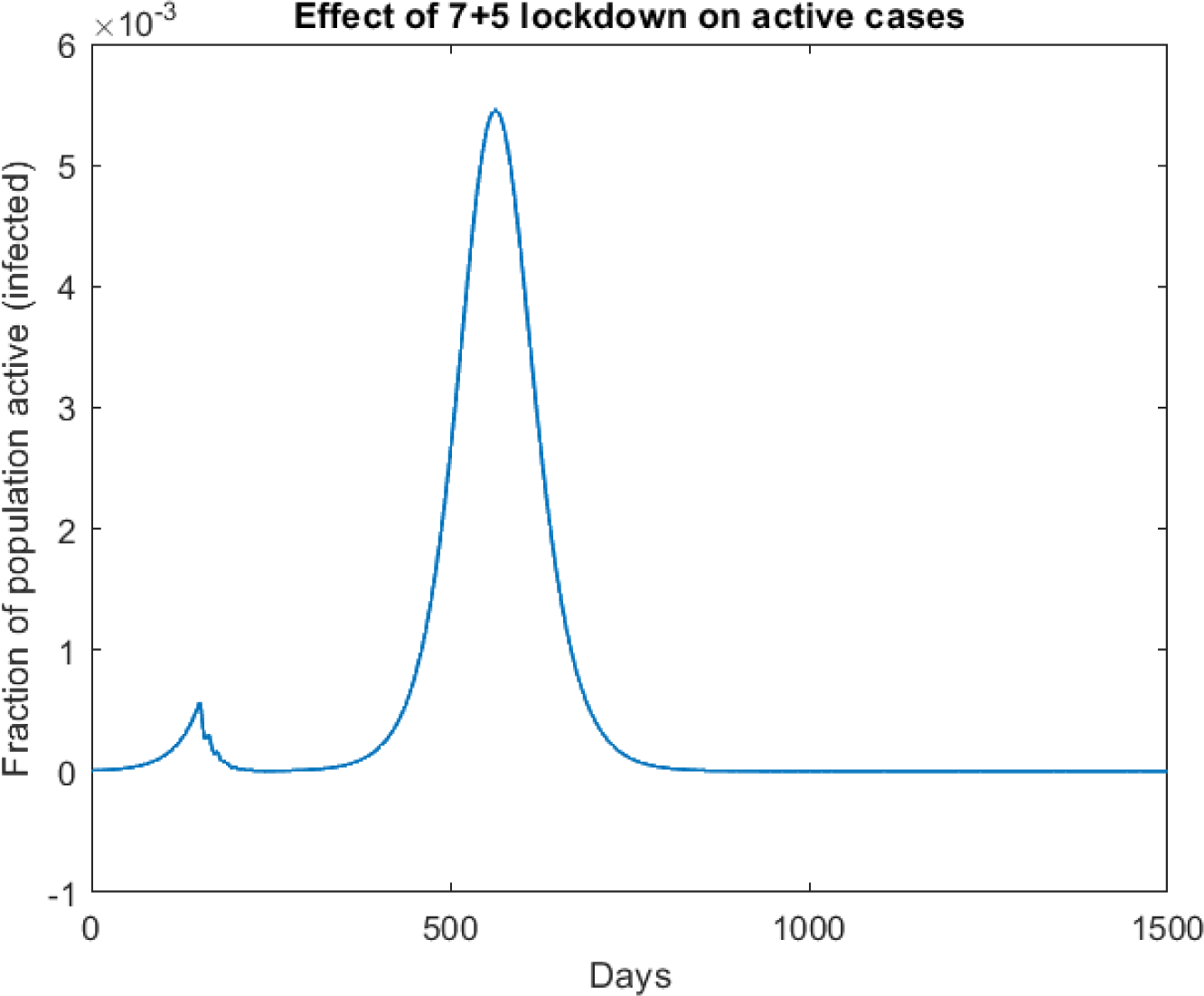
7+5 lock down for 150-250 days

#### 5.4.2 Light-switch lock down

Another strategy that can be employed is to lock the economy down when the number of reported cases cross a certain threshold. Since in our model, the only cases that the government can document is of those displaying symptoms or those currently admitted in a medical facility, we have simulated scenarios in which light-switch lock down has been employed when the the net values of *I_s_ + Q + Q′* cross a pre-determined threshold value. The simulations for the active cases at different threshold levels are depicted in Fig. 11.

From the plots given in Fig. 11, it is evident that light-switch lock down tends to stretch out the period of the epidemic. Lower the threshold, longer the duration. The number of cases in this model too is constant. But unlike the periodic or continuous lock down models, this gives an advantage to the system. The longer duration of the epidemic implies that region’s medical system receives sufficient time to recover from the extra load it had to face, during the onset of the epidemic. Thereafter, since the number of cases never increases beyond a certain value, by controlling the threshold, we can ensure that most patients get adequate health care.

**Assumptions made:**

- The entire system under consideration is locked down simultaneously at the point of enforcement and the lock down is relaxed at every region of the system simultaneously, when done so.
- At least one individual remains infectious after the lock down is relaxed.
- No violations of lock down occur during the entire period the lock down is imposed.
- No migration from other cities/systems occurs to the system under consideration.

#### Recommendations for public policy

The manner in which the lock downs should be imposed by the government should be based on basic game theory rules. Locking down the economy would yield a decrease in the spread. But keeping the economy shut for too long can lead to economic depressions and increased filings for bankruptcies and unemployment which has to be seriously considered before deciding the extent of the lock down. So, the fashion in which the lock down policies would be implemented should be such, that the utility gained during the entire time period of the epidemic by keeping the economy is optimized, while not allowing the epidemic to go out of control.

**Figure 11:**
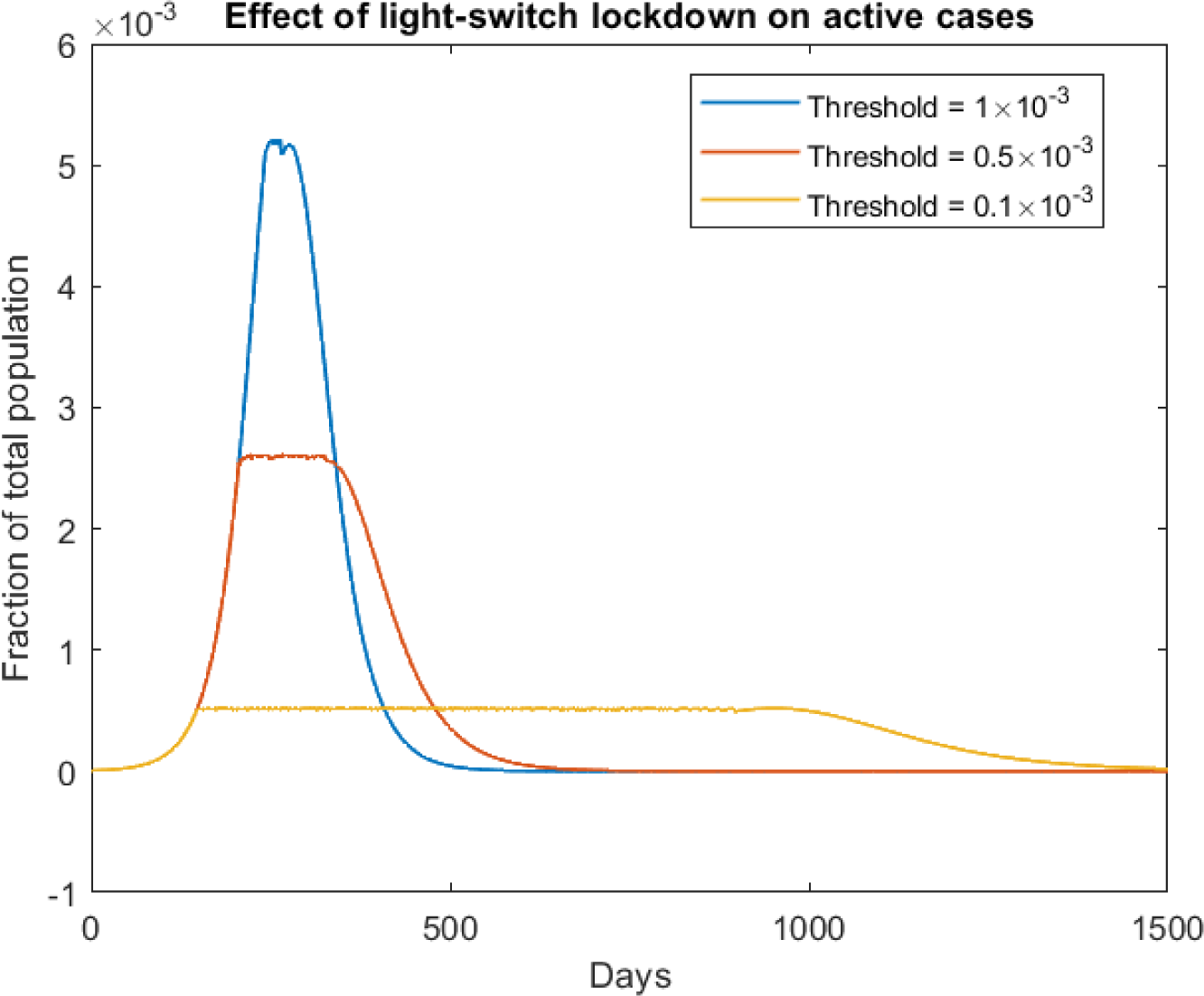
Effect of light-switch lock down on active cases

#### 5.4.3 Violation of lock down

As depicted in several of our previous simulations, locking down can lead to a delay in the peak of the epidemic, which can prove favourable for the government to set up crucial facilities needed for fighting the epidemic. However, a point we did not elaborate on, is what if lock down is not followed religiously. In a real sense, it is very much probable for a section of individuals to violate lock down due to personal reasons. To take this into account, instead of defining *α* to a definite value at the time of lock down (0.14 in our previous simulations), we may define it as a periodic function oscillating between a definitive maximum and minimum values with the period referring to the time scale with which lock down is violated periodically. For the purposes of our simulation, we have used

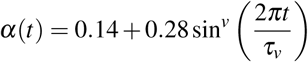
where *v = 2n*, 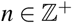 governs the spread of the violation cycle (higher the value, more condensed the spread) and *τ_v_* gives the time scale of period of the cycle. The coefficients 0.14 and 0.28, indicate that the oscillation of *α*(*t*) is within 0.14 and 0.42 (our original post-lock down and pre-lock down values). As will be apparent soon, it is the spread that is a major deciding factor in how the dynamics of the epidemic and even a minimal violation of lock down will cause the delay time (caused by lock down) to be reduced significantly. Fig. 12 depicts the simulations if lock down is imposed for days 150-200 for varying values of *τ_v_* with *v* fixed at *v =* 2.

**Figure 12:**
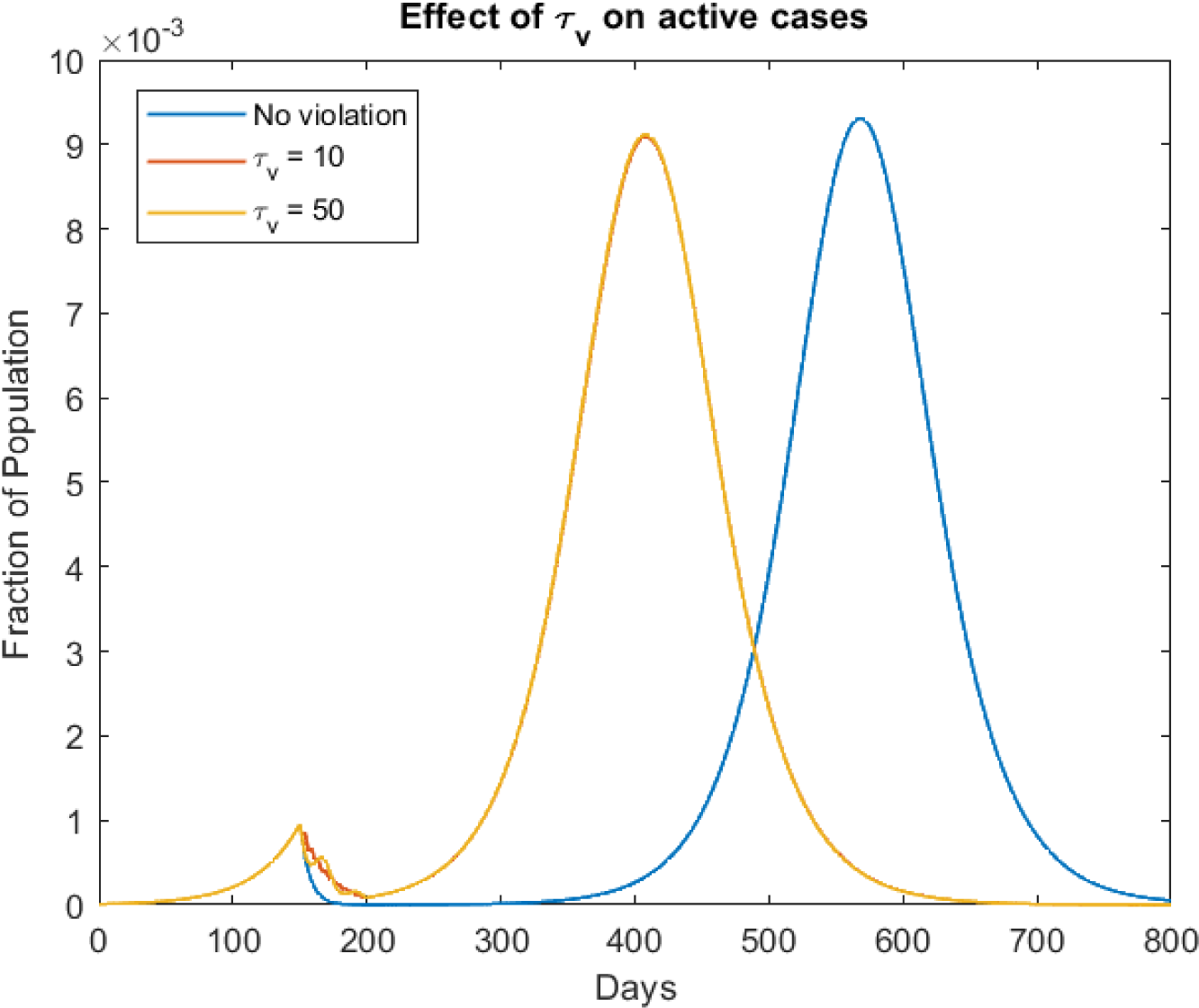
Effect on the total active cases: *I_s_ + I_as_ + Q + C* for v = 2

**Figure 13:**
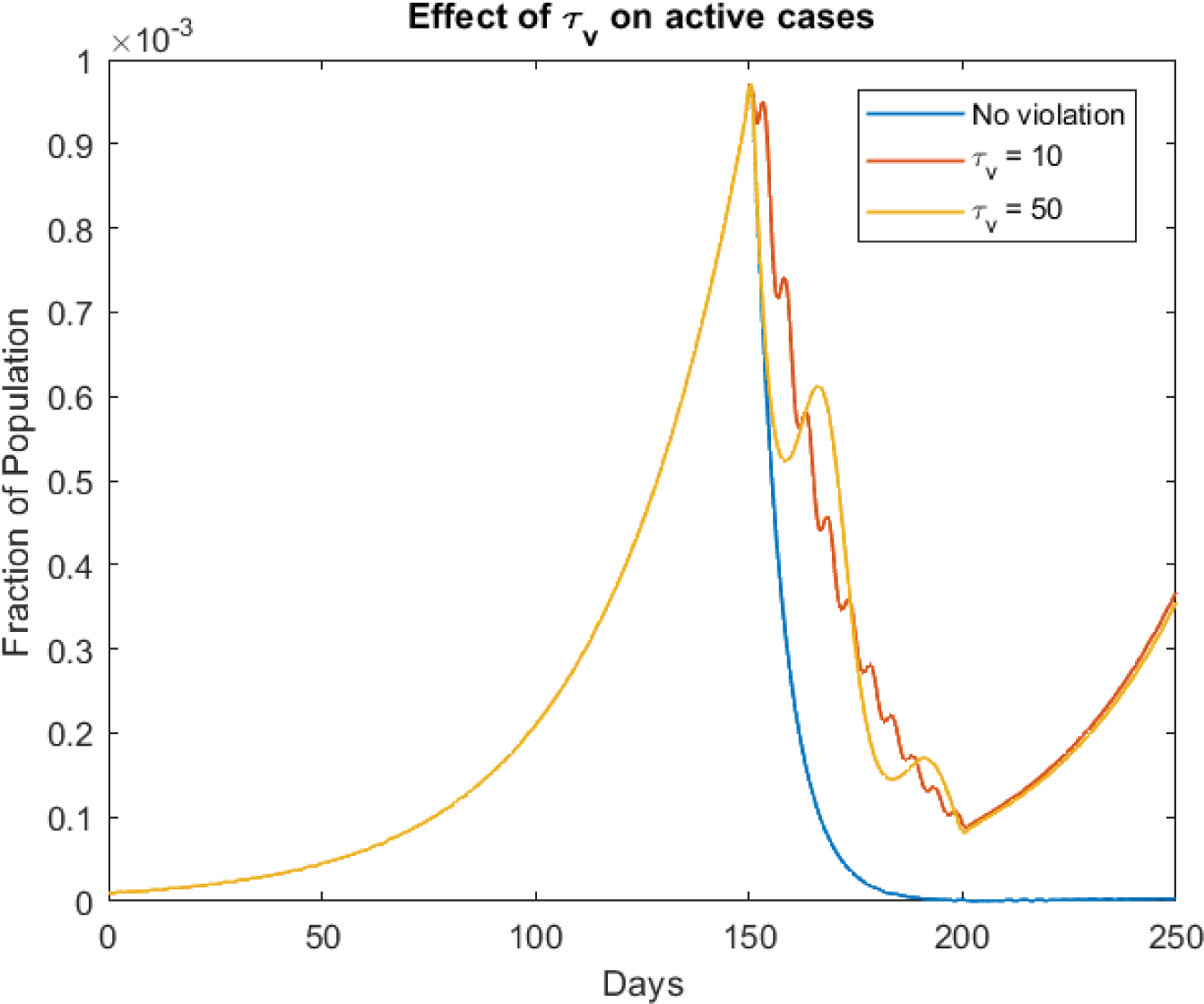
A zoomed picture of the above plot for t<250

From the above figures, it is evident that events of violation of lock down will diminish the delay time (between first wave and second wave) that could have otherwise proved vital for helping the medical system recover. As mentioned earlier, it is the spread that is the major factor that affects the dynamics of the epidemic and not the time period of the cycle. Fixing *τ_v_ =* 10, Fig. 14 is the simulation for varying values of v, if lock down is imposed for days 150-200.

**Figure 14:**
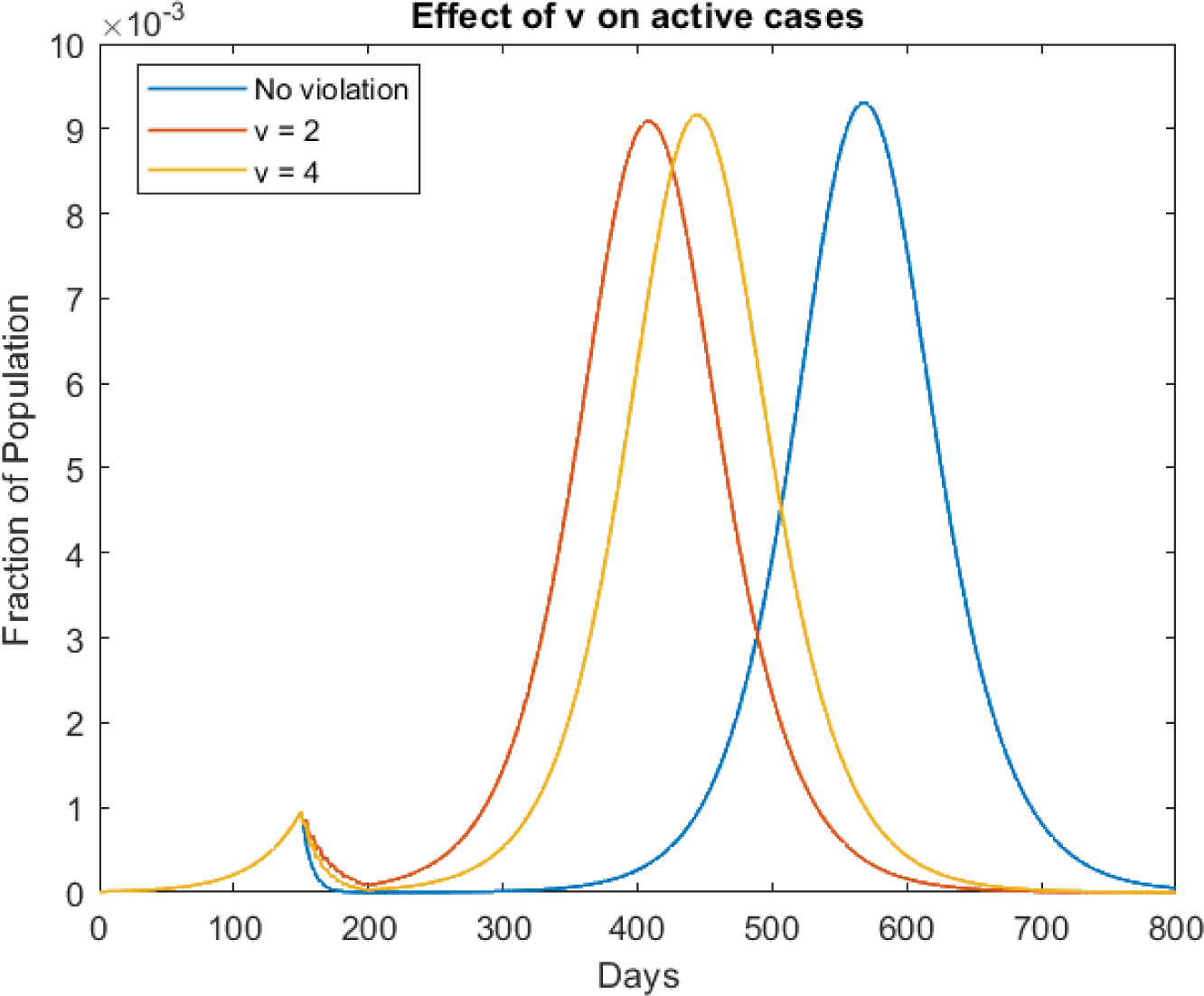
Effect on the total active cases: *I_s_ + I_as_ + Q + Q′ + C* for *τ_v_ =* 10

As seen in the above plot, a more condensed violation (higher value of v) would help the delay go on for a longer time. The no violation case, in fact corresponds to the limiting case, when *v* tends to infinity.

**Assumptions made:**

- The entire system under consideration is locked down simultaneously at the point of enforcement and the lock down is relaxed at every region of the system simultaneously, when done so.
- At least one individual remains infectious after the lock down is relaxed.
- The violations of lock down occur in a periodic fashion.
- No migration from other cities/systems occurs to the system under consideration.

#### Recommendations for public policy

From the above results, it may be possible that a periodic lock down works better. Since the guarantee of the lock down being lifted in the near future, may decrease violations (thus condensing the spread), so that the delay time is not diminished significantly.

### 5.5 Setting up quarantine and health-care facilities

In our model, the parameter *ε* determines the rate with which symptomatic infected individuals who have approached a medical health-care facility requesting treatment are being quarantined. As mentioned in section 3.1, this was introduced to account for the possibility of a collapse in medical infrastructure in case of an uncontrollable surge in the number of confirmed cases. For the simulations done by far, we have assumed it to be constant at *ε =* 0.5. But, if the government takes appropriate measures to begin construction of medical facilities to curb the epidemic, the story will be totally different. In this subsection, we will analyse what happens if instead of a constant, *ε* is an increasing function of time (*t*). For this purpose, we have used the function:

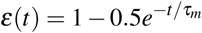

where *τ_m_* corresponds to the time-scale of increase in the value of *ε*. Observe that at *ε*(0) = 0.5, which is the value we used in our previous simulations, and lim*_t_*_→∞_*ε*(*t*) = 1. The simulations for various values of *τ_m_* are depicted in Fig.15 and 16.

**Figure 15:**
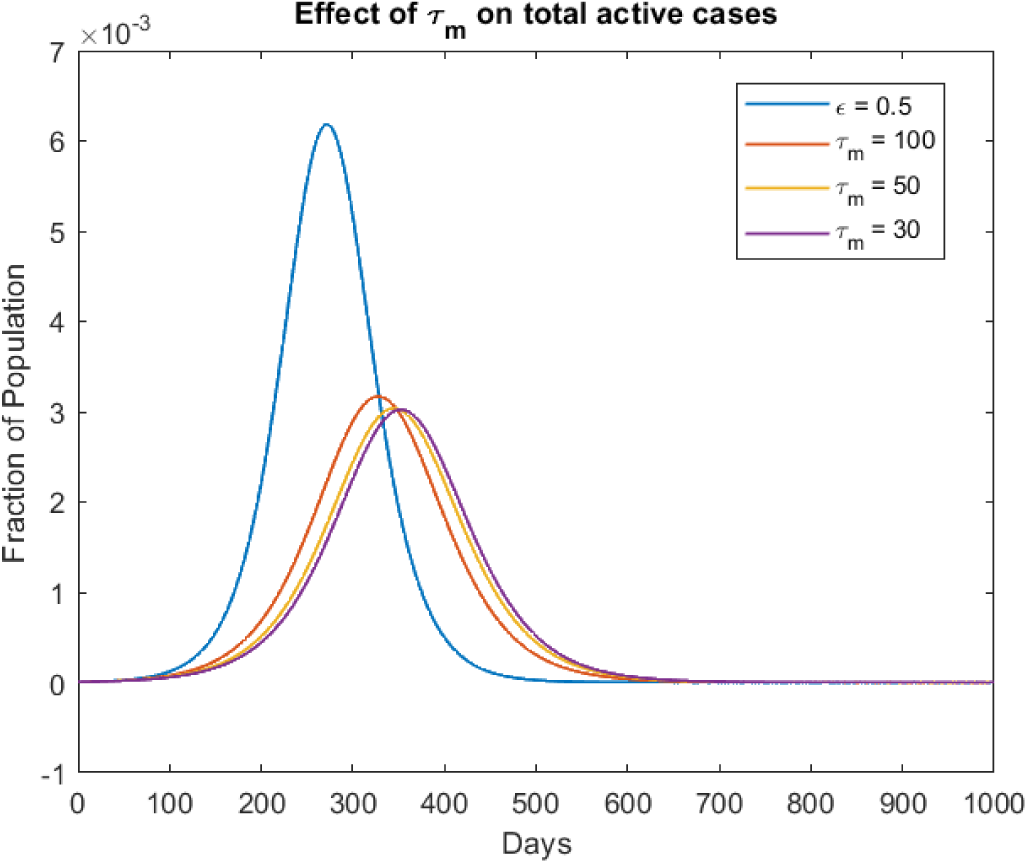
Effect on the total active cases: *I_s_ + I_as_ + Q + Q′ + C*

**Figure 16:**
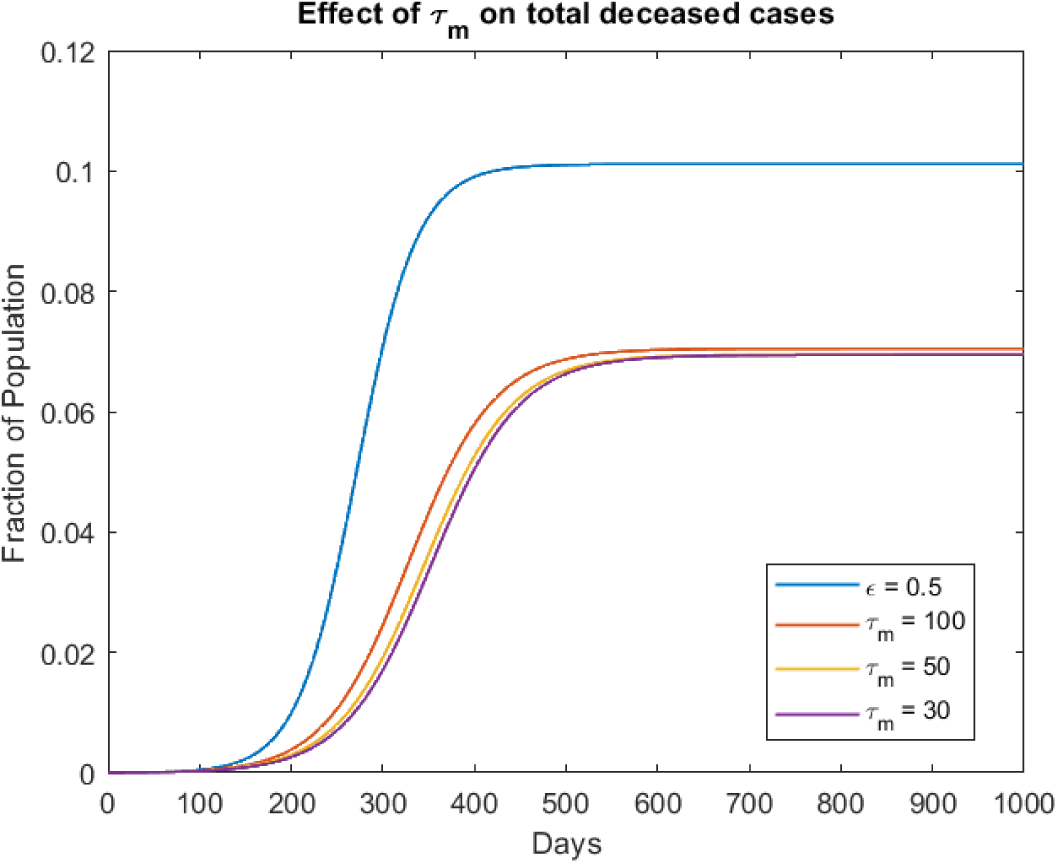
Effect of total deceased cases: D

**Assumptions made:**

- The health-care systems are set up in equal proportions at every region of the system.
- There is no discrepancy of accessing health-care facilities at two different locations in the system under consideration.
- No lock down is imposed.
- No migration from other cities/systems occurs to the system under consideration.

#### Recommendations for public policy

As observed in the above plots, the spread of the infection is dramatically reduced as *τ_m_* decreases. So, the government should take systematic and stringent measures to set up health-care and quarantine facilities around the country, since this would decrease the number of cases of infection significantly

### 5.6 Strategical quarantining and testing

In all of the simulations done by far, it is only symptomatic individuals who are quarantined and thereby given the appropriate treatment in a medical facility. Asymptomatic individuals are never identified. Thus it seems that the bulk of the infections are spread by the *I_as_* compartment. This issue can only be solved if strategic testing and quarantining is done on mass populations so as to identify such silent carriers and quarantine them. To incorporate this feature in our model, the equations for the two concerned compartments *I_as_* and *Q* have to be slightly modified. The modified equations are as follows:

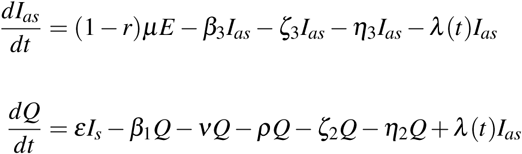

where *λ*(*t*) is the identification rate of asymptomatic individuals. Now, before applying this testing strategy, it is important for the ruling body (government) to first set up sufficient medical/quarantining facilities and ensure that the symptomatic individuals, who have approached the hospital are all accommodated in some health-care facility. Else, enacting policies to test asymptomatic individuals before ensuring the treatment of the cases at hand does not make sense. Hence, this idea has to be used along with the model described in the section 5.5. From now, we set *ε*(*t*) *= 1 − 0.5e^−t/τm^* with *τ_m_ =* 100 and

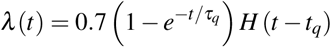

where *τ_q_* determines the time-scale with which the efficiency of detection of asymptomatic cases increases and *H*(*t*) is the Heaviside step function. This has been included to take care of the delay of implementation of this testing and quarantining strategy. *t_q_* is the parameter which takes care of this delay time. In Fig.17 are the simulations for various values of *τ_q_* keeping *t_q_ =* 200.

**Figure 17:**
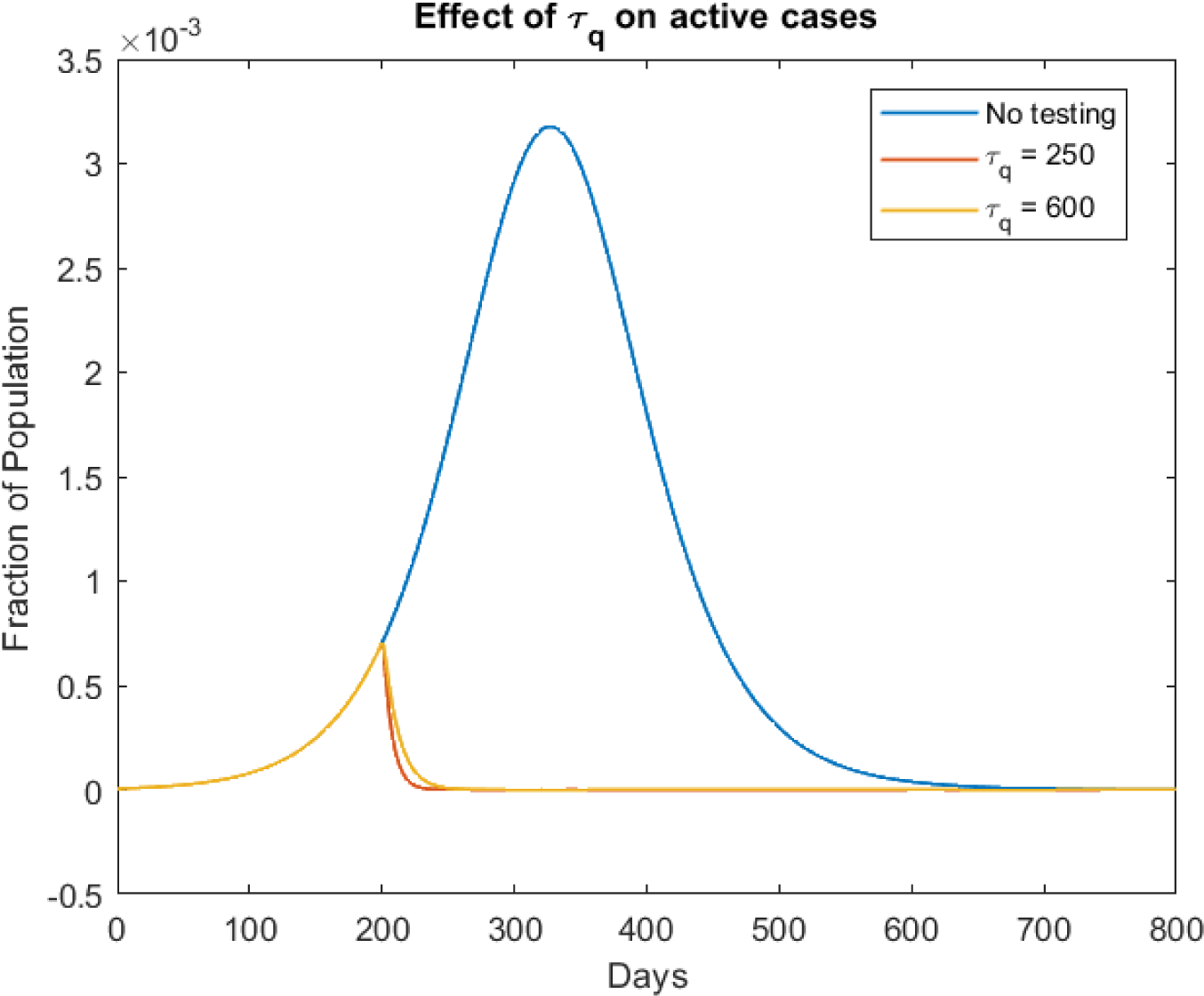
Effect on the total active cases: *I_s_ + I_as_ + Q + C* for *t_q_ =* 200

**Figure 18:**
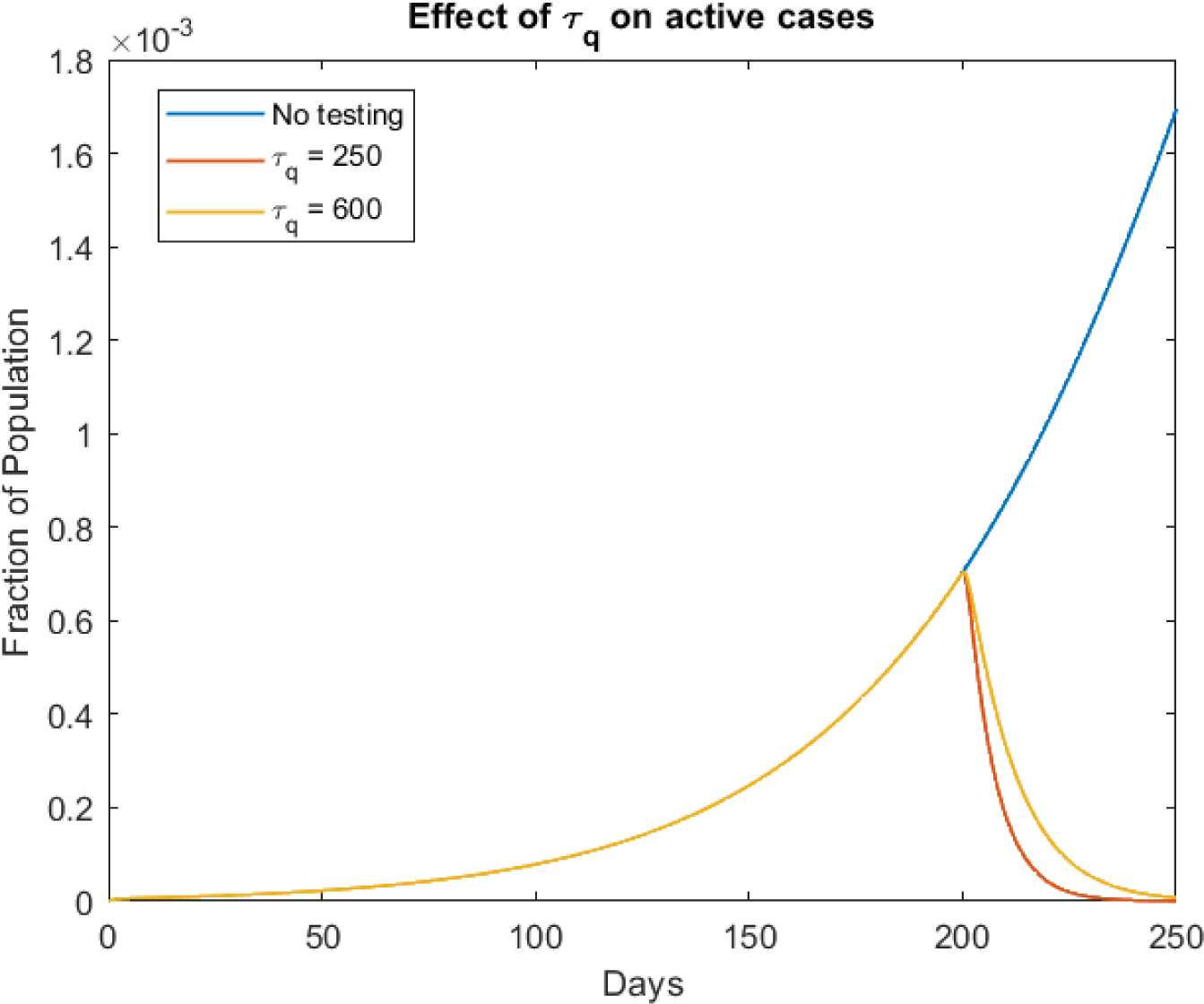
A zoomed picture of the above plot for t<250

From the above figures, it is evident that a serious employment of the testing and quarantining strategy can decrease the number of active cases manifold. Longer it takes for the government to employ this strategy, the more it can cost in terms of the number of people under its care. The simulations in Fig.19 will make this point clear, where we have simulated the effect on active cases for varying values of *t_q_* keeping *τ_q_ =* 250.

**Figure 19:**
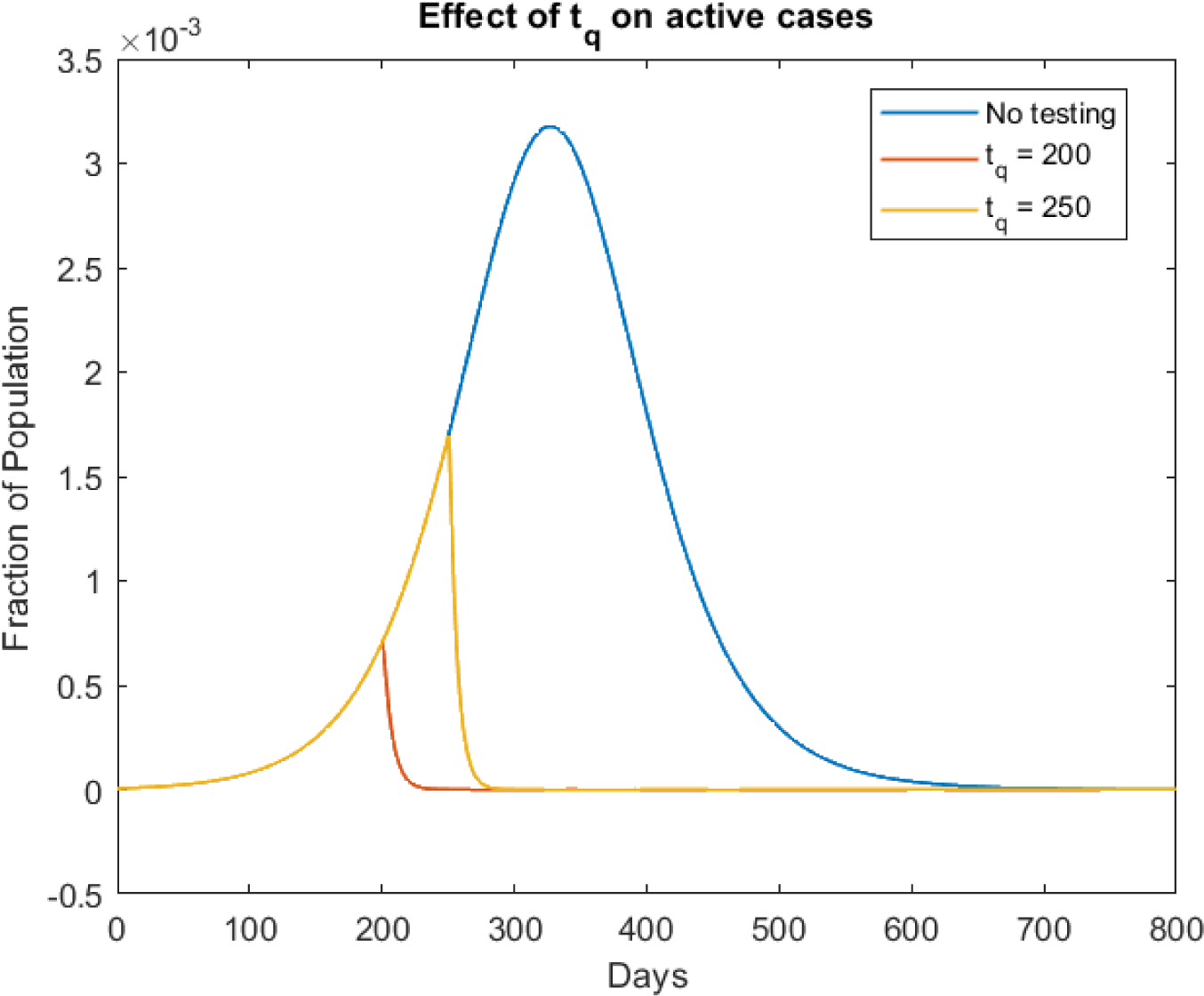
Effect on the total active cases: *I_s_ + I_as_ + Q + C* for *τ_q_ =* 250

As seen in the above plot, the sooner is this strategy employed (lesser *t_q_*), the faster will this epidemic get over.

**Assumptions made:**

- The health-care systems are set up in equal proportions at every region of the system.
- There is no discrepancy of accessing health-care facilities at two different locations in the system under consideration.
- Testing is not done as age/travel history specific.
- No lock down is imposed.
- No migration from other cities/systems occurs to the system under consideration.

#### Recommendations for public policy

From the above result, it is clear that testing and quarantining of asymptomatic individuals is a crucial step that the government should take to decrease the number of cases. Taking into account the highly contagious nature of COVID-19, implementing this soon will not be easy, since the number of reported cases itself may overwhelm the medical infrastructure. So, it is advised that as soon as the threat of the pandemic is detected, immediate steps should be taken to set up more hospitals and quarantine facilities. Thereafter, mass testing and quarantining of detected positive cases should be done substantially.

### 5.7 Effect of migration

In this subsection, we will look at the effect of immigration on the economy and try to visualize how the infection spreads to different regions over time. For now, we will only consider the migration due to compartments *S, I_as_* and *E*. This is based on the assumption that the other compartments comprise of those who have contracted the virus and are aware of it, hence would ideally, avoid travel.

The modified differential equations are as follows:

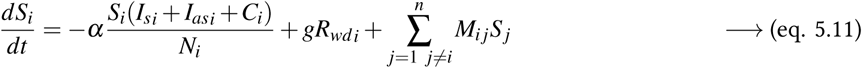

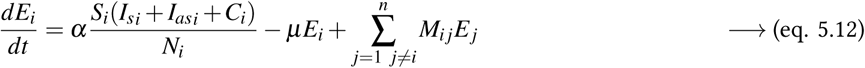

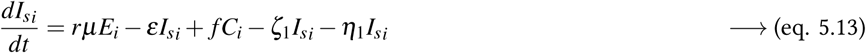

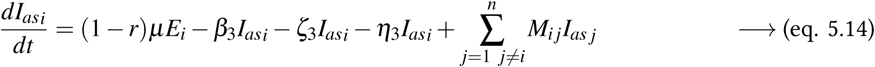

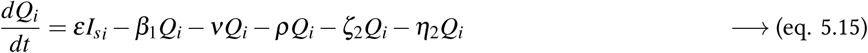

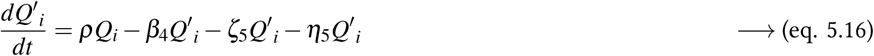

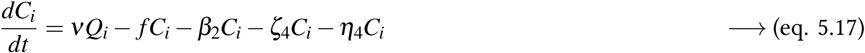

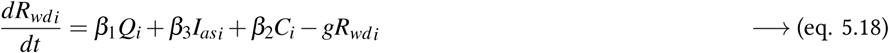

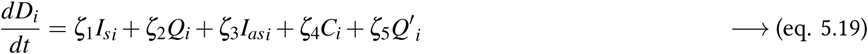

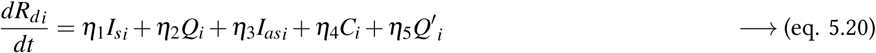

where 1 ≤ *i, j ≤ n* corresponds to the variables for the *n* locations being considered. Here *i* corresponds to the target location (to where immigration happens) and *j* corresponds to the source location (from where emigration occurs). For simplicity we assume that the rate-coefficients do not depend the geographical region index.

#### 5.7.1 Deciding *M_ij_*

The migration coefficient should have the following properties:

1. In the earlier days of the epidemic, when reports of infections begin to float on media, it is natural for people to panic, thereby resulting in an initial surge in the number of migrations because of speculations of an imminent lock down. Thereafter, this number will decline, on general awareness to avoid travel at times of emergency.
2. It should be a characteristic function of the existing economic conditions of *i* and *j*.

From property 1, it is evident that we cannot define *M_ij_* as a constant, instead it will be a function of time. Keeping both properties in mind, the form of *M_ij_* can be displayed as *f*(*t*) × *π_i,j_* where *f*(*t*) corresponds to the functional form taking care of property 1. A good choice for *f*(*t* is:

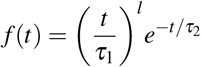

where τ_1_ determines the peak value of the migration rate and *τ*_2_ and *l* determines the time scale with which the migration numbers reaches its maxima ad subsequently dies out. *l* also determines the rate of the surge in migration.

Now, at any instant, the total number of individuals in each of the compartments *S, I_as_* and *E* due to migration is conserved i.e.

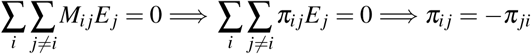

#### 5.7.2 Simulating effect of migration on our model

The anti-symmetric matrix [*π*]*_ij_* chosen for this simulation is given below:

|  | City 1 | City 2 | City 3 | City 4 |
| --- | --- | --- | --- | --- |
| City 1 | 0 | -5 | -10 | -15 |
| City 2 | 5 | 0 | 7 | -10 |
| City 3 | 10 | -7 | 0 | -5 |
| City 4 | 15 | 10 | 5 | 0 |

Taking *τ*_1_ *=* 10, *τ*_2_ *=* 3 and *l* = 4, the simulation for a system of 4 cities is done in Fig. 20, when the first patient (patient zero) belongs to city 1.

The higher number of cases in City 4 as seen from Fig. 20 is no coincidence. Observe that *π_ij_* is positive for *i =* 4∀*j* ∈ {1,2,3,4}. This indicates that maximum people migrated to City 4, hence the relatively higher number of cases. Similarly, maximum people migrated from City 1, hence it is the least hit location. Also, it seems that the first location to become free of the pandemic is the one where patient zero was located (City 1). Note that in these simulations we have considered the population of each of the compartments to be equal. More interesting behaviours will be observed if adding more details to this migration model.

**Figure 20:**
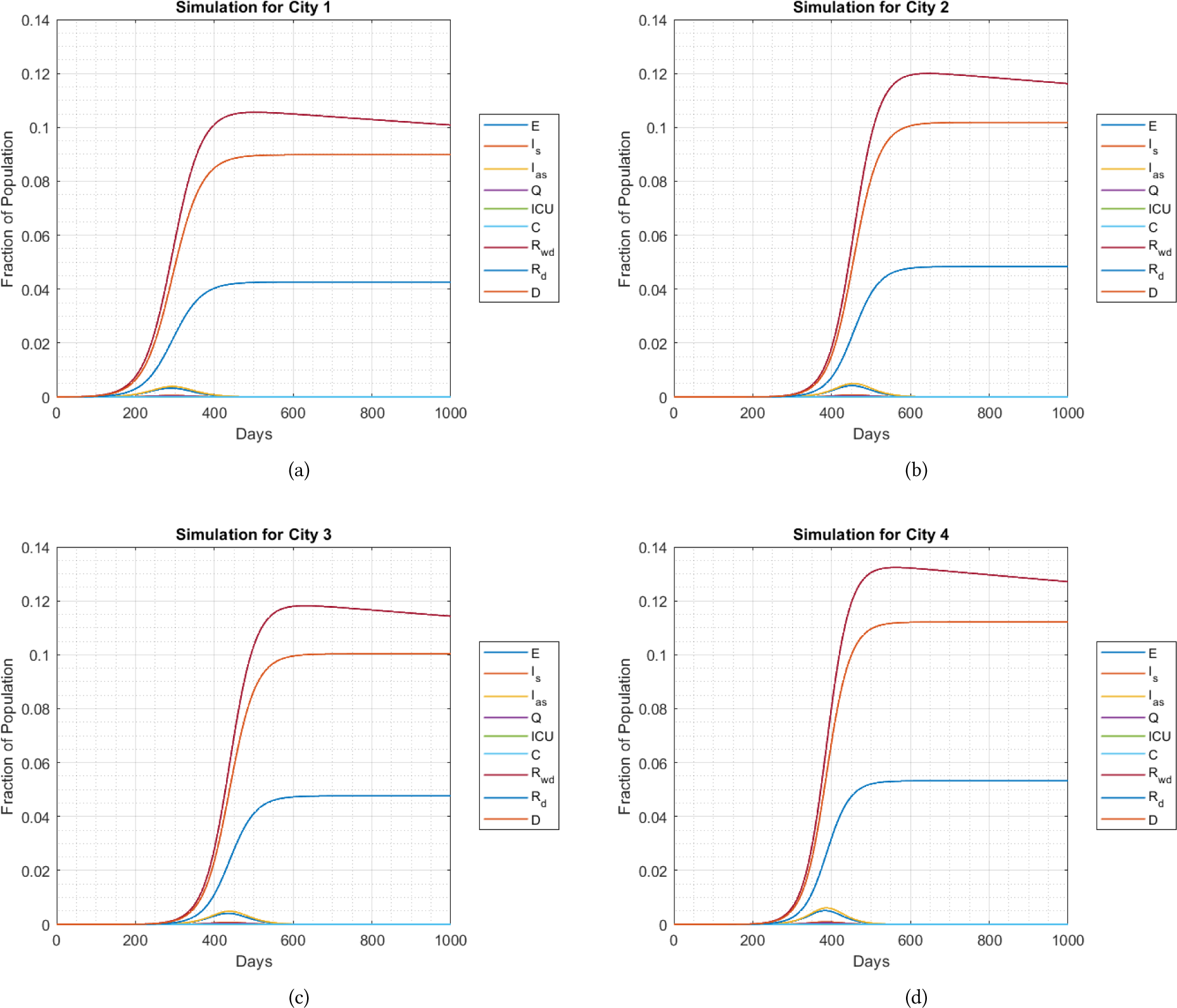
Patient zero belongs to City1

## 6 Comparing simulations with real documented data

In this section, we will compare some simulations with the actual data of active cases of India, obtained from authentic sources such as [John Hopskins CSSE](https://github.com/CSSEGISandData/COVID-19) + [fixes](https://mackuba.eu/corona/overrides.json?1587060786) data set and www.covid19.org

As observed in all of our previous plots, just imposing a lock down is sufficient to decease the number of cases to near zero. But the actual trend of active cases does not follow this pattern. The plot of the active cases in India from January 30, 2020 (the day patient zero was identified) to April 26, 2020 is given in Figure 21.

**Figure 21:**
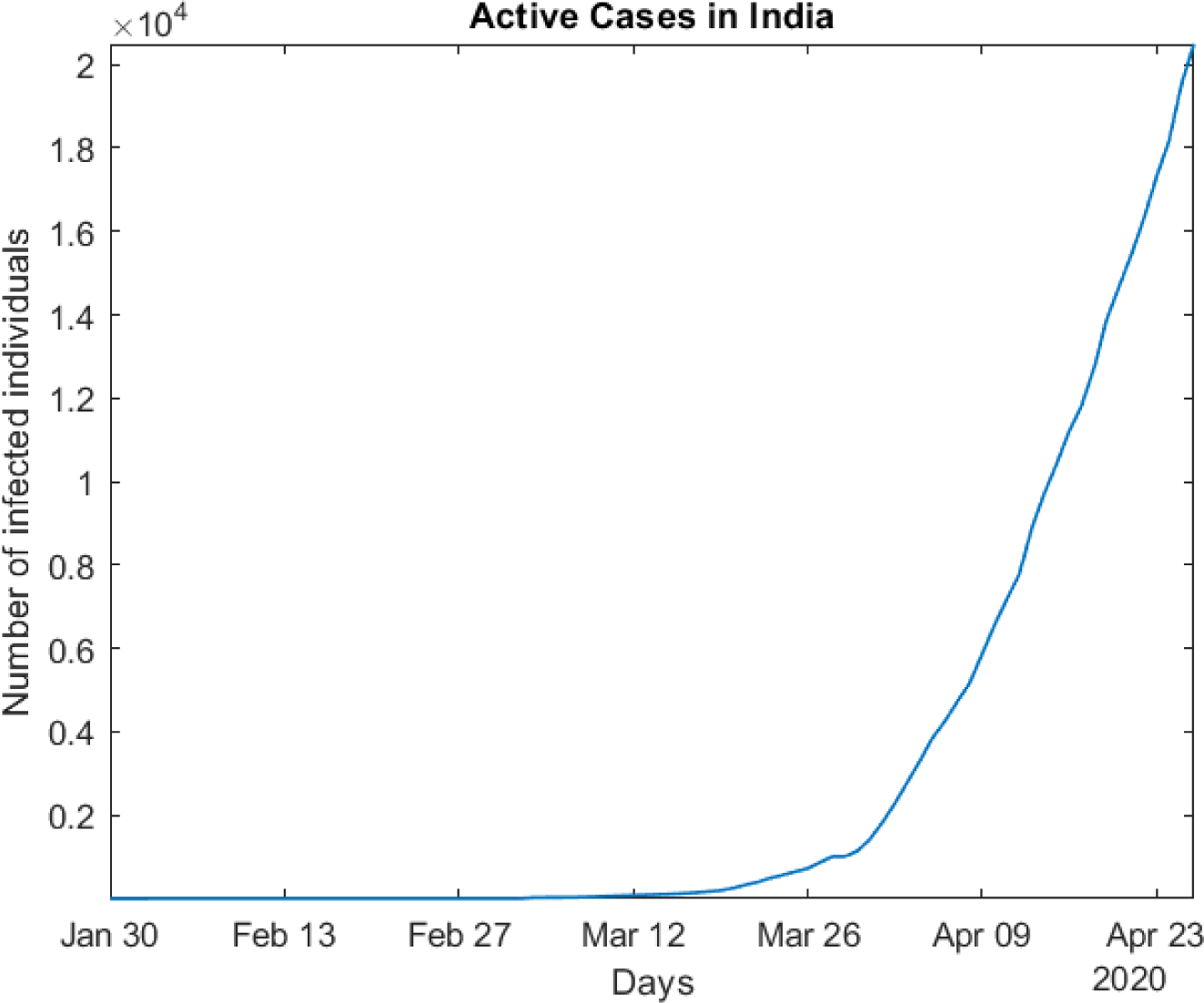
Active Cases timeline of India

This anomaly may have occurred because of a number of reasons. Three major discrepancies we identified are as follows:

1. In our model we have used the *I_s_* compartment to accommodate all those individuals who developed symptoms and subsequently visited a medical facility to acquire treatment. But in doing so, we have assumed that all the individuals who develop symptoms in the economy will seek treatment without haste. But in real world, this may not necessarily be true. It is highly likely that some individuals opt to remain silent on the issue to avoid social distancing, quarantine and other measures that may be against his personal interests.
2. In our model, we have defined the parameter *ε* as the rate with which symptomatic individuals are quarantined. In addition to assuming the simultaneity in contracting the infection and reporting it to the hospital for the individual in *I_s_* compartment, we also assumed that the test results are hundred percent accurate. However, this is not necessarily true. Researchers from the UK’s University of Bristol claim that as high as [30 percent of false negatives are generated in the current test of COVID-19](https://www.weforum.org/agenda/2020/04/coronavirus-covid19-tests-pandemic-virus-antibodies-swab-blood/). So, it is a real possibility that some symptomatic individuals are not able to avail quarantine because they were wrongly tested negative, and hence are unwillingly infecting other susceptible individuals they come in contact with.
3. In our previous simulations, we assumed that the act of imposing a lock down would immediately be followed by a decrease in the value of the parameter *α*. But this is not necessarily true. On speculations of a lock down, some days prior to it, there will be a rush among the public to get back to their residences before the lock down begins. This increases the interaction rate between people. The hike will continue for a few days after the lock down is imposed and thereafter *α* will decrease and attain a minimal value.
4. As mentioned in section 5.3.3, some of the parametric values were borrowed from the INDSCI-SIM study. It turns out that their model is quite different from ours not only in terms of the equations, but also in terms of the compartments used. So, the same parametric values might not necessarily hold in our model.

To account for the point 1 in the list of discrepancies, we have come up with a small change in our compartmental model. We’ve introduced a new compartment called reluctant symptomatic 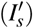 to accommodate those symptomatic individuals who have ignored their symptoms and have thus not approached a health-care facility yet. Note that eventually, these symptomatic individuals are going to wilfully go to the hospitals when then condition worsens. This is why the 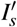 compartment is not same as the *I_as_* compartment, where the individuals would never go to quarantine (*Q*) unless explicitly tested by the testing and quarantining strategy discussed in section 5.6. Our modified compartmental model looks like the following:

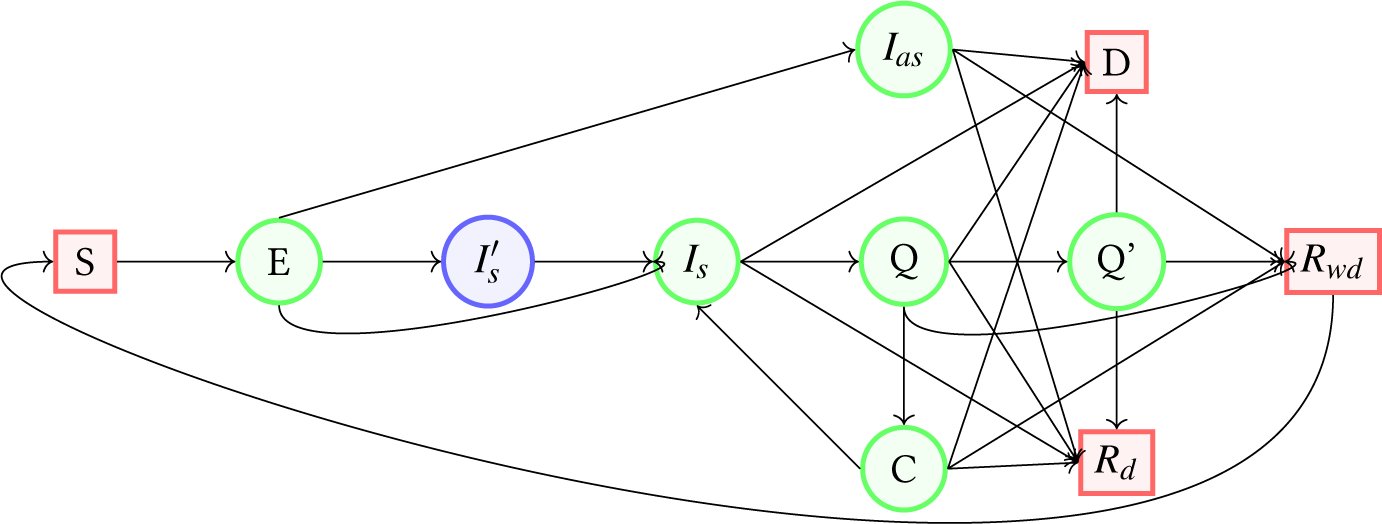

All the equations stated in equations 5.01 - 5.10 is unchanged except that for 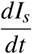. Also a new equation for 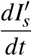 has to be introduced. The modified differential equations for these compartments are as follows:

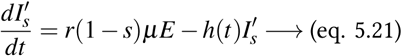

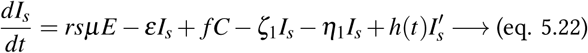

where *s* is a ratio which measures how much proportion of the individuals developing symptoms do not immediately visit the hospital. *h*(*t*) is a time-dependent function which determines the time-delay the individuals in 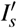 compartment exhibits before visiting a health-care facility and thereby get quarantined.

Intuitively 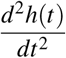 will be positive for some *t < τ_w_*. Thereafter, it will be negative. The following choice for the functional form of *h*(*t*) can been made:

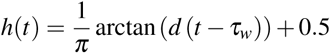

*τ_w_* determines the initial value of *h*(*t*). It also gives a measure of the time delayed by individuals in 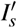 on an average before visiting the hospital. *d* gives a measure of the distribution of *h*(*t*). This parameter is defined to take into account the asynchronism of time delay between different individuals in 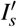.

As mentioned in point 2, in the list of discrepancies, the parameter *ε* needs to be taken care of to accommodate for false negatives. To solve this issue, we have changed the definition of some of our parameters and compartments a bit. Initially in section 5.5, we had attributed *ε* to the simply the rate with which more medical facilities are build across the nation. But, *ε* should also account for the medical efficiency (accuracy of the tests). Now, taking into account the early stringent lock down measures that the Government of India has taken, the accuracy factor seems to be a far more dominant factor of the two. So, we redefine *ε*(*t*) as the rate which which the accuracy in COVID-19 test is increased, thereby giving lesser false negatives with time. Subsequently we redefine *τ_m_* (used in section 5.5) to the measure of the time-scale with which this is achieved.

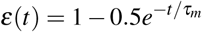

Note that we have not changed the coefficients 1 and 0.5, because coincidentally it gives a good estimate of the initial testing accuracy (0.5) and the testing accuracy which we wish to achieve after a long time (1.0).

Since we have changed the definition of *ε*(*t*), now, there would be some symptomatic individuals who approached the medical facility and were tested negative. But, they are still able to infect others. Now, numerous possibilities arise for such individuals :

1. They fall sick at a later time, approach the hospital are tested positive and are thereby quarantined.
2. They fall sick later, approach the hospital, but are again negative negative (wrongly) and are again left. This process can be repeated many times unless the individual is either quarantined or the loop is terminated (the individual never visits a hospital after certain iterations).
3. They never approach a hospital again.

Our initial model cannot explain such transitions. So, we have decided to change the definition of our *I_as_* and *I_s_* compartments. We define the compartment *I′* as all those individuals who are infected, but gets quarantined (unless explicitly made to if caught by the testing and quarantining strategy) before either recovering or dying or recovering with permanent disability. The individuals in this compartment may be symptomatic (but tested negative) or asymptomatic. We define another compartment *I* as all those individuals who would are infected and would necessarily eventually get quarantined wilfully (not by being found positive in the testing and quarantining strategy). Now *I_as_* in our previous model will be replaced with *I′* and *I_s_* will be replaced with *I*. Note that the difference between *I′* and 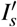 (the new compartment that was added to take care of individuals who are delaying to report their symptoms to the hospital) is that individuals in *I* would report their symptoms immediately, while those in 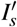 would not. Both these compartments contain individuals who will necessarily be quarantined eventually by their own will. Our modified compartmental model looks like the following:

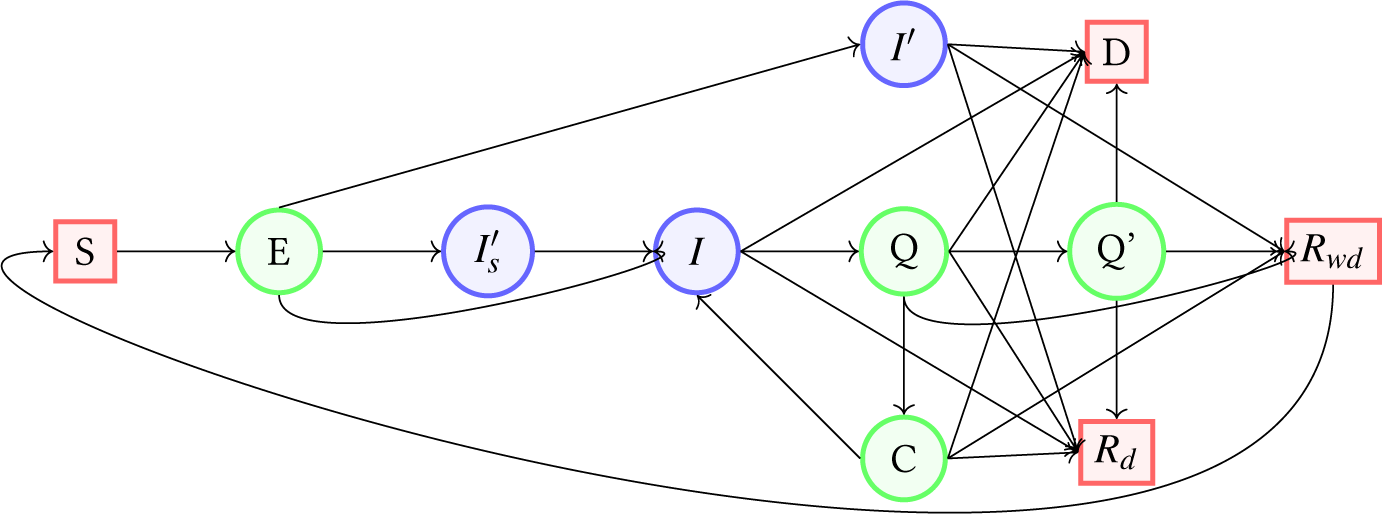

Another change which we mentioned was necessary was the nature of the variation in the parameter *α* just before and immediately after lock down is imposed. It will substantially increase some days prior to lock down and will begin to decrease some days after lock down. Keeping this nature in mind, the following functional form has been set for *α*.

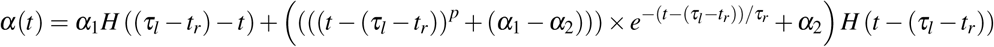

where *τ_l_* is the time after which lock down is imposed. *t_r_* is the measure of the time before lock down when the individuals in the system begin to rush back to their residence. *τ_r_* is the time scale with which *α*(*t*) dies down after lock down. *α*_1_ is the value of lock down before *t = τ_l_ − t_r_* i.e. before the rush occurs and *α*_2_ is the minimum value of *α*(*t*) that is reached some time after lock down is imposed. *H*(*t*) is the Heaviside step function.

Since April 2, 2020, the government started strategic testing and quarantining of asymptomatic individuals. Hence, to get an effective simulation we would have to use the results obtained in section 5.6 as well.

The set of differential equations being used to run our simulation now looks as follows:

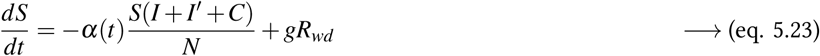

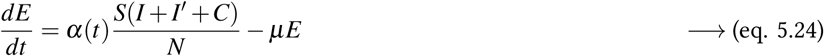

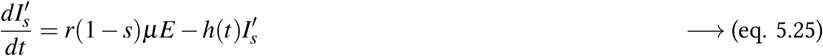

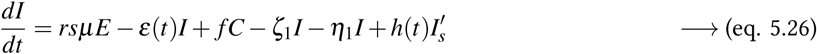

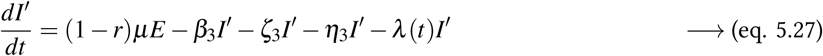

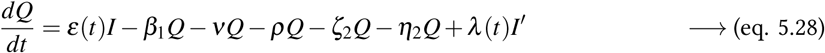

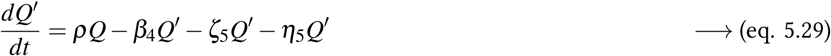

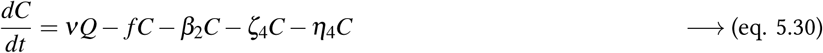

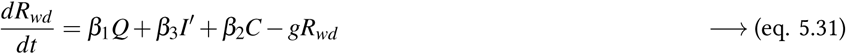

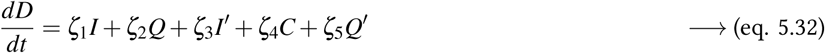

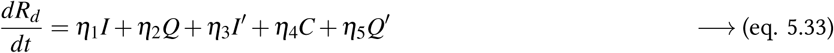

where the functional forms of the time dependent parameters are as follows:

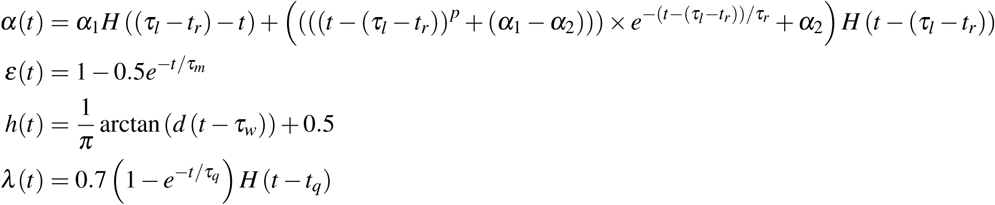

As mentioned earlier, the values of parameters we have used in our previous simulations do not give expected results. So the next simulation has been done after setting the parameters studying the literature survey of COVID-19 pandemic and estimating good values that fit to the definition of parameters used for India. The table of parameters used are as follows:

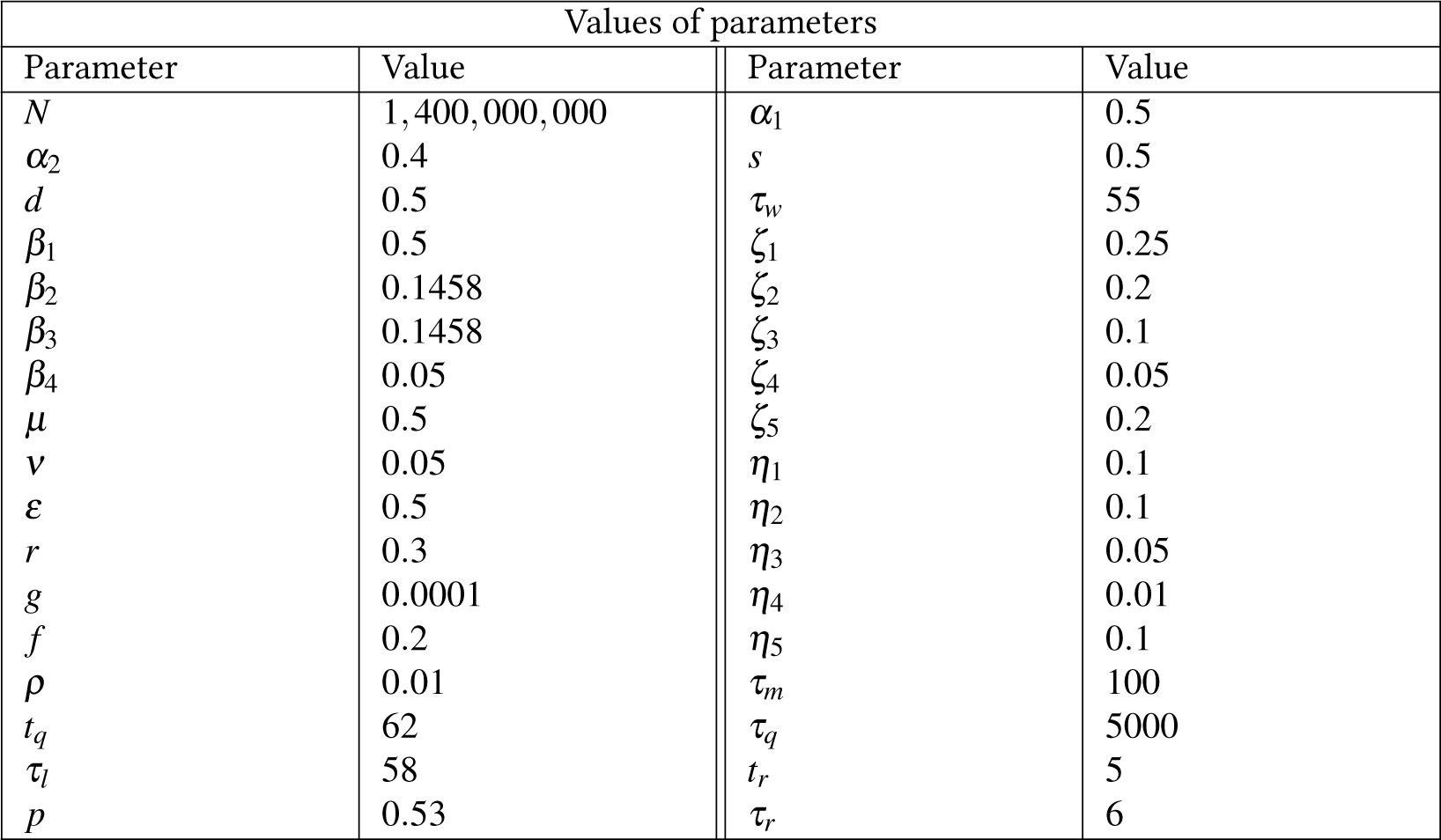

The simulation obtained by our model is compared with the actual documented data below:

**Figure 22:**
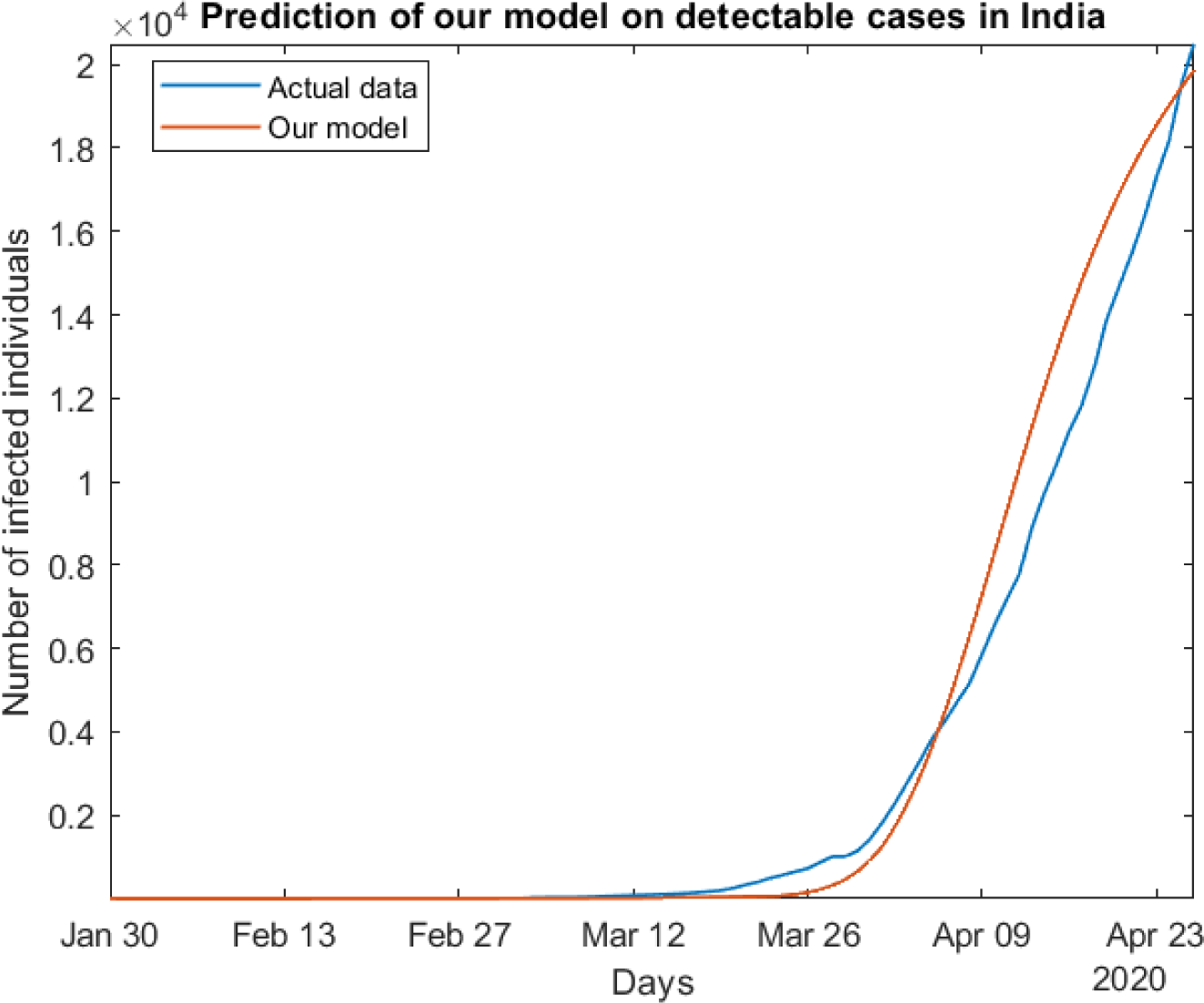
Comparison with actual data. *N*(*0*) = 1399999964, *E*(0) = 25, 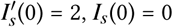; *Ias*(*0*) = 2, *Q*(0) = 2, rest = 0

## 7 Highlights

1. **Excluding all other external effects, will imposing a lock down lead to a decrease in number of cases?** A — No. If we exclude key factors like strategic testing, setting up of medical facilities, etc., imposing a lock down will not decrease the number of cases. It will simply delay the onset of the pandemic. The total number of cases would still remain the same.
2. **Why should the lock down be imposed in the first place if it does not decrease infections?** A — Imposing a lock down will give a temporary relief by decreasing the number of cases. This delay time can be utilized to do important things like setting up medical infrastructure, increase the rate of testing and quarantining, etc., so that the next wave that will follow would not be as lethal as it would be if no steps are taken.
3. **Will the number of infections remain low if the lock down is lifted in near future?** A — Any brief lock downs will only delay the onset of the peak of the pandemic. If regular testing and quarantining is done, the number of infections will decrease if the lock down period is suitably long.
4. **What will happen if lock down is lifted on 6 July, 2020 (100 days after lock down was imposed)?** A — Upon lifting lock down on this date, the pandemic could spin out of control - unless rigorous testing and quarantining of infected individuals is done before lifting the lock down. Our model estimates that 14.6 crores of individuals could die and 41.33 crores individuals (more than two-fifths of the population) could be affected.
5. **What will happen if lock down is lifted on day 22 January, 2021 (300 days after lock down was imposed)?** A — In this case, the pandemic could still lead to devastating results. Our model estimates that 7.03 crores individuals could die and 19.79 crores individuals (almost one-seventh of the population) could be affected (unless testing and quarantining of infected individuals is done more rigorously before lifting the lock down).
6. **How should the lock down be imposed to minimise the death count?** A — Imposing lock down until the disease disappears would be the most optimal solution. In our model, the number of individuals in each compartment can take fractional values like 0.6, 1.3, etc. In reality, such values represent the integers closest to them, since all calculations should be in natural numbers. So, if the number of infections is less than unity, it would mean that there are no active cases. So imposing lock down till number of infections becomes less than unity will amount to the least number of deaths.
7. **Will the number of infections become zero if 1 year of lock down is imposed?** A — No, even though one year of lock down may seem excessive, it is still not sufficient to clear away all the infections from India (unless testing and quarantining of infected individuals is done more rigorously before lifting the lock down).
8. **How long should the lock down be imposed in India for the number of infections to become zero?** A — Assuming no significant violations of lock down occur, India would have to remain locked down till day 732 (lock down was imposed on day 58) to get rid of all infections (assuming the testing and quarantining procedures are done at the current rate), thus ending the pandemic.
9. **How many total deaths would occur if lock down is imposed till day 732? What will be the peak value of active cases in a day? How many of these will be detected? How many individuals will be affected during the pandemic?** A — Assuming that the virus does not mutate and no vaccine is found, and we largely forgo testing and quarantining procedures, 38.8 lakh deaths would occur due to COVID-19 in India. The peak value of active cases in a day would be 1.53 lakh. Of these 0.29 lakh would be detected. The pandemic would affect a total of 1.8 crore individuals.
10. **Why are cases in India still increasing after lock down is imposed?** A — There can be multiple reasons for this. Even though lock down has been imposed, it is not strictly followed. There are several places where proper care is not being taken. Violation of lock down means that temporary curbing of the pandemic does not occur. So the number of cases will continue to rise. Also testing and quarantining of the infected remains very poor in India.
11. **How can we decrease the number of cases?** A — The most effective solution is to increase the rate of conducting tests. Asymptomatic individuals have to be identified as quickly as possible to curb the pandemic.
12. **What happens if disease transmission rate** (*α*) **is negative?** A — If this is the case, then the number of infections quickly die out and the pandemic would be over soon. With the initial conditions considered for the result obtained in section 5.3.4, the number of infections died out in roughly 25 days for *α =* −0.01. This result agrees qualitatively with the findings in the paper [COVID-19 in India: State-wise Analysis and Prediction](https://www.medrxiv.org/content/10.1101/2020.04.24.20077792v1), authored by Palash Ghosh, Rik Ghosh and Bibhas Chakraborty.

**Note:** The above results are what our deterministic model predicts which is subject to the simplifying assumptions made while deriving them. Actual numbers may be different in case stochasticity is important, which the present study ignores. Future deleterious mutations of COVID-19 may exacerbate the situation whereas development of an effective vaccine may mitigate the problem.

## 8 Conclusions

This project proposal includes substantial new developments including the derivation of Compartment Models that are specific to COVID-19 which includes rates of death, disability and quarantine numbers which the simpler models do not include. In the model, we also distinguish between asymptomatic/symptomatic infections. These newly derived models have been solved and results displayed. Further developments will involve using machine learning and other tools to find a scientific way of pinning down adjustable constants in the models. A more thorough study of the effects of inter-state migration on the epidemic is still pending. Lastly, effects of stochasticity have to be thoroughly studied.

## Data Availability

With regards to the authenticity of the paper, we would like to state that the ideas we have presented in the paper are our own unless otherwise explicitly specified. And we have, to the best of my knowledge, mentioned the sources of the concept, data or assertion whenever they were borrowed. In doing so, we have provided due credit for any work conducted by other individuals, groups or institutions.

## Appendix

### INDSCI-SIM

[A state-level epidemiological model for India: INDSCI-SIM](https://indscicov.in/for-scientists-healthcare-professionals/mathematical-modelling/indscisim/?fbclid=IwAR21o9WgQoL0yZIgI4JOWznV7f7XlPrW9GOuu9WgT823TCqoPtD1hCPlFmM)

Snehal Shekatkar, Bhalchandra Pujari, Mihir Arjunwadkar, Dhiraj Kumar Hazra, Pinaki Chaudhuri, Sitabhra Sinha, Gautam I Menon, Anupama Sharma, Vishwesha Guttal

INDSCI-SIM is the first detailed, state-specific, epidemiological compartmental model for COVID-19 in India.

**Objectives**

- Their work include the following objectives:
- To develop an India-specific state-level model to predict spread of COVID-19 using state-of-the-art epidemiological models.
- Calibrate model to clinical parameters, so that these can be used as benchmark numbers for modeling.
- Incorporate state-specific demographic data.
- Model different non-pharmaceutical interventions (NPI) strategies in each state.
- Make all the codes and online tools available.
- Flexible enough for easy update and changes transparently communicated.

**Similarities with our model**

- It features both the susceptible and exposed compartment in the same definition in our model.
- Exactly two compartments arise from the exposed compartment (into pre-symptomatic and asymptomatic), like our model (asymptomatic and symptomatic). The parameter *γ* used in this work is analogous to the parameter used *r* used in our work.
- Only symptomatic individuals are hospitalized.
- Unlike some famous fundamental models like SIR, SEIR, etc. and alike our model, this work too have separated the deceased compartment from the recovered compartment.

**Dissimilarities from our model**

- This work have introduced compartments like pre-symptomatic and mildly symptomatic, which we have clubbed together with the symptomatic and asymptomatic compartments in our model.
- In our model, we have included a compartment for those people who have recovered from the disease, but in the process have incurred permanent disability, unlike this work.
- In our model, we have distinguished between patients who are mildly and critically ill by segregating the total hospitalized in the Quarantined (*Q*) and ICU (*Q′*) compartments.
- In our model, we have introduced a carrier state compartment (*C*) to account for inefficiency in medical checking, wherein some individuals may be wrongly tested negative and thus subsequently, leave quarantine.
- In this work, an individual can die only after being hospitalized. We have relaxed this requirement in our model, wherein transitions can occur to the deceased (*D*) or disabled (*R_d_*) compartments from any of the following compartments: Symptomatic infected (*I_s_*), Asymptomatic Infected (*I_as_*), Quarantined (*Q*), ICU (*Q*) and Carrier State (*C*).
- In our model, we have accounted for the possibility of re-infection (after subsequent loss of immunity upon recovery) by incorporating a transition line from Recovered (*R*) compartment to the Susceptible (*S*) compartment.

### Works of some Epidemiologists

- **Anthony Fauci** *Director of the National Institute of Allergy and Infectious Diseases.*

– [Coronavirus Infections—More Than Just the Common Cold](https://jamanetwork.com/journals/jama/article-abstract/2759815) In this article, he discusses • Why Corona virus infections are much more than just common cold • The History of SARS dating back to 2002 • The symptoms and mortality rate • Difference between MERS and SARS • Countermeasures being taken against Covid-19
– [Covid-19 — Navigating the Uncharted](https://www.nejm.org/doi/full/10.1056/nejme2002387) In this article, he discusses • Median age of Patients (59) • Age factor in the mortality rate • Effect of Asymptomatic cases on the mortality rate • Government strategies and Vaccines
– [Novel vaccine technologies for the 21st century](https://www.nature.com/articles/s41577-019-0243-3) In this article, he discusses • Novel approaches to vaccine development • key insights that enabled the production of a stabilized subunit vaccine candidate • Technical advances in mRNA vaccines • revaccination of uninfected adolescents
- **Jeffrey Shaman** *Infectious disease expert at Columbia University.*

– [Substantial undocumented infection facilitates the rapid dissemination of novel coronavirus (SARS-CoV2)](https://science.sciencemag.org/content/early/2020/03/24/science.abb3221) In this article, he discusses • The effect of undocumented infections on dissemination • A mathematical model that simulates the spatiotemporal dynamics • time-to-event observation model using a Gamma distribution • Application of model-inference framework to the observed outbreak
– [Initial Simulation of SARS-CoV2 Spread and Intervention Effects in the Continental US](https://www.medrxiv.org/content/10.1101/2020.03.21.20040303v2) In this article, he discusses • Use of metapopulation model applied at county resolution to simulate the spread and growth of COVID-19 • Projection of the outbreak in the continental US for 180 days after March 13 • The effects of social distancing and travel restrictions on the outbreak
– [Direct Measurement of Rates of Asymptomatic Infection and Clinical Care-Seeking for Seasonal Coronavirus](https://www.medrxiv.org/content/10.1101/2020.01.30.20019612v1) In this article, he discusses • Rates of Asymptomatic Infection • Clinical Care-Seeking for Seasonal Coronavirus • findings from a proactive longitudinal sampling study
- **Gerardo Chowell** *Mathematical epidemiologist, Georgia State University.*

– Estimating the asymptomatic proportion of coronavirus disease 2019 (COVID-19) cases on board the Diamond Princess cruise ship, Yokohama, Japan, 2020 In this article, he discusses • Cases on board the Diamond Princess cruise ship, Yokohama, Japan • statistical modelling analysis to estimate the proportion of asymptomatic individuals • Laboratory testing by PCR
– [Transmission potential and severity of COVID-19 in South Korea](https://www.ijidonline.com/article/S1201-9712(20)30150-8/fulltext) In this article, he discusses • Transmission potential and severity of COVID-19 in South Korea • The mean reproduction number of COVID-19 in Korea • fatality rate is higher among males and increases with age.
– [Estimating Risk for Death from 2019 Novel Coronavirus Disease, China, January-February 2020](https://www.ncbi.nlm.nih.gov/pubmed/32168464) In this article, he discusses • Estimating Risk for Death from 2019 Novel Coronavirus Disease, • time-delay adjusted risk for death from COVID-19 • breakdown of the healthcare system • enhanced public health interventions, including social distancing and movement restrictions.
- **Juan Gutierrez** *Mathematics proffesor, University of Texas at San Antonio.*

– [Investigating the Impact of Asymptomatic Carriers on COVID-19 Transmission](https://www.medrxiv.org/content/10.1101/2020.03.18.20037994v3) In this article, he discusses • Impact of Asymptomatic Carriers on COVID-19 Transmission • Inaccuracy of current Reproduction number • effective reproduction number could range from 5.5 to 25.4 • agreement with average case data collected from thirteen countries
– [An Epidemiological Model of Malaria Accounting for Asymptomatic Carriers](https://www.ncbi.nlm.nih.gov/pubmed/32172448) In this article, he discusses • Asymptomatic individuals in the context of malarial disease • Rigorous mathematical analysis of a new compartmentalized malaria model accounting for asymptomatic human hosts • qualitative analysis will fill in the gaps of what is currently known
- **Tara Smith** *Epidemiologist, Kent State University.*

– [Report from the American Society for Microbiology COVID-19 International Summit, 23 March 2020: Value of Diagnostic Testing for SARS-CoV-2/COVID-19](https://mbio.asm.org/content/11/2/e00722-20) She discusses about • Value of Diagnostic Testing for SARS-CoV-2/COVID-19 • types of tests available and how they might be useful • Test for Viral RNA • Serology

^1^

## Footnotes

Atul Kumar was initially in the list of authors of this project. After he left the project to pursue his personal interests vested in study of high energy physics, his name was dropped from the list of authors. Anish Chandrachud was added to the list of authors after he expressed his interest towards this project and is now an active contributor.

